# Modelling a tobacco-free generation policy in Australia: population health impacts under illicit market uncertainty

**DOI:** 10.64898/2026.07.08.26357588

**Authors:** Samantha Howe, Tim Wilson, Coral Gartner, Tony Blakely, Driss Ait Ouakrim

**Affiliations:** Melbourne School of Population and Global Health, University of Melbourne; School of Public Health, University of Queensland

## Abstract

**Objective:** To estimate the potential health and equity impacts of a tobacco free generation (TFG) and T21 policy (increasing the legal age of sale to 21) in Australia, in the context of a complex market including widespread illicit tobacco and e-cigarette product availability.

**Design:** A Markov macrosimulation model, parameterised with yearly net movements between legal smoking, illicit smoking, vaping, and dual use states, combined with a proportional multi-state lifetable.

**Setting:** The Australian population, modelled as an open cohort for 40-years.

**Intervention:** A ‘business-as-usual’ (BAU) scenario was compared to TFG and T21 policies, with both starting in 2026. Variations to policy impacts were tested under increasing background illicit market enforcement.

**Main outcome measures:** The model estimates the health-adjusted life years (HALYs) and deaths over 40 years, under each scenario, with differences across age and socioeconomic status (SES) presented.

**Results:** The TFG policy reduced daily smoking prevalence among 15-24-year-olds to 5.3% (95% uncertainty interval [UI] 4.8-6.0%) in 20 years’ time, compared to 7% under the T21 policy and 7.6% under BAU trends. Vaping was minimally impacted by either policy. The TFG policy resulted in 75,000 (95% UI 41,900-124,000) HALYs being gained over 40 years. The policy impact was larger when accompanied by increased illicit market enforcement, reducing daily smoking among 15-24-year-olds to 1·3% within 20 years. Both policies had greater prevalence and health impacts on more disadvantaged compared to advantaged SES groups.

**Conclusion:** A TFG policy is expected to produce long-term benefits for the Australian population but would be most effective in combination with increased enforcement of illicit tobacco and e-cigarette markets. Novel strategies to increase quitting in addition to reducing uptake are needed to improve tobacco-related outcomes in the short to medium term.

**Plain language summary:** *The known:* A ‘tobacco free generation (TFG)’ policy would theoretically minimise youth smoking uptake in Australia but may be undermined by rising illicit tobacco and e-cigarette use.

*The new:* A TFG law in Australia may reduce daily smoking among 15-24-year-olds by 30% within 20 years, though resulting health impacts take longer to manifest. Larger impacts would also occur if smoking uptake via the illicit market was well controlled.

*The implications:* A TFG policy could produce long-term benefits in Australia but should be implemented alongside policies to reduce illicit product supply and improve smoking cessation among adults who already smoke.

## Introduction

Ending commercial tobacco use requires policies that both support cessation among people who currently smoke and prevent future cohorts from becoming addicted. The tobacco free generation (TFG) proposal, first described in 2010, takes a cohort-based approach to tobacco sales: people born after a specified year would never become legally eligible to buy tobacco products [1]. This differs from a Tobacco 21 (T21) policy, which delays legal purchase until age 21. A TFG policy therefore has a broader goal of changing societal norms, based on the argument that new cohorts should not be legally permitted to enter lifelong addiction to a harmful product.

Interest in age- and birth-cohort-based tobacco policies has increased internationally. T21 was adopted federally in the United States in 2019 [2] and Ireland in 2024 (commencing in 2028 [3]), and the UK has now legislated a generational tobacco sales ban, expected to commence in 2027 [4]. In Australia, no generation-based tobacco ban is currently in place. However, 2024 federal legislation banned all e-cigarettes from general retail sale for all ages, limiting legal access to pharmacies who supply for smoking cessation purposes [5,6].

The potential impact of a TFG policy in Australia is uncertain, as the tobacco and wider nicotine market is changing rapidly. Regular e-cigarettes use among 15-24-year-olds increased to 17% in 2022-23 [7], and the majority of e-cigarette uptake appears to occur via illicit market access^8^ despite recent legislative changes. Further, an increasing proportion of tobacco purchasing occurs via the illicit market in Australia, with recent estimates suggesting that illicit tobacco comprised 50-60% of total tobacco consumption in 2024-25 [8]. The effect of a TFG policy could be blunted if included cohorts can readily access illicit tobacco or e-cigarettes.

Simulation modelling has estimated the potential health benefits of age- and cohort-based tobacco policies in several settings [9–12], but Australian evidence remains limited. Existing models also provide limited insight into how illicit tobacco and e-cigarette availability may modify policy impact. In this study, we use the SHINE-Tobacco simulation platform to estimate the population health and equity impacts of a TFG policy in Australia, compared with business-as-usual trends and a T21 policy. We explicitly model legal and illicit tobacco use, vaping and dual use, and assess how policy impacts vary under alternative levels of illicit tobacco and e-cigarette market enforcement.

## Methods

### Overview

An existing macrosimulation modelling platform, SHINE-Tobacco [12–14], is used for this analysis. The platform is comprised of a discrete time Markov process, used to simulate yearly changes to smoking and vaping behaviours [14], and a Proportional Multi-State Lifetable (PMSLT) [15,16], which tallies the impacts of changing smoking/vaping rates on 31 tobacco-related diseases in the Australian population. Each component of the model is disaggregated across sex, age, geographic remoteness, Indigenous status, and socioeconomic status (using Socio-Economic Index For Areas [SEIFA] quintiles [17]), as described in detail in Howe et al. [13]. For this analysis, we focus on the output across age groups and SEIFA quintiles.

The business-as-usual (BAU) model was constructed based on historic epidemiological data, assuming trends in disease rates, all-cause mortality rates, and overall smoking prevalence will continue.[13] The inputs of the BAU model are shown in Supplementary Table 1.

Two main intervention scenarios, representing a TFG and T21 policy, were modelled on top of the BAU scenario, both in isolation and in combination with increased illicit tobacco and e-cigarette market enforcement.

### Business-as-usual model

#### Markov model

The Markov model incorporates yearly net transitions between daily smoking, vaping, and dual use of both products. To form the BAU scenario, smoking uptake and quit rates were first fit to existing daily smoking prevalence forecasts by sociodemographic group [18]. Vaping and dual use rates were then calibrated to recent prevalence by sociodemographic group [7], with two separate states defined for vaping: ‘naïve’ vaping arising from no previous product use (accessed via illicit market), and vaping as a behaviour post-smoking (including a combination of illicit market and pharmacy access). Smoking and dual use uptake and quit rates, and smoking intensity, were then split into values for illicit vs. legal states based on the difference in price of these products (at a price elasticity of −0·4 [19]). The rate of switching from legal to illicit smoking was calibrated to achieve an increase in the proportion of smoking that is via illicit supply from 25% in 2023 to 50% in 2025.[8,20] As a sensitivity analysis, the modelled interventions (described below) were also compared to variations in the BAU Markov model, produced in a previous analysis by varying the switching transitions between smoking and vaping. Further details are provided in the Supplementary Material, and in previous work (Howe et al. [14]).

#### PMSLT

The PMSLT models the Australian population as an open cohort for 40-years, in a lifetable defined by births, migration, mortality rates, and morbidity rates [21]. Disease lifetables are defined by forecast incidence, remission and case fatality rates by cohort, based on historic trends [22], and static disability weights. The forecast parameters were held constant after 20 years.

Changes in the distribution of the population in the Markov model states under an intervention flow through to changes in disease incidence via a population impact fraction (PIF). The PIF uses disease incidence rate ratios (RRs) that follow dose-response curves based on smoking intensity (in cigarettes per day or pack-years), estimated for each sociodemographic group [23]. Given the lack of existing data on long-term vaping-related health risk, we made the following assumptions. First, e-cigarettes smoked per day was assumed to match that for tobacco smoking for the same sociodemographic group. Second, the disease incidence RRs for each smoking-related disease were scaled down, assuming a mean 89% (95% UI 80-95%) lower risk for vaping compared an equivalent amount of cigarette smoking. Scaling to 60% lower risk than smoking was used in a sensitivity analysis. These ranges reflect that applied in previous modelling [24,25]. Disease RRs in the former use states take 30 years to decline to no excess risk. Resulting changes in disease morbidity and mortality are summed to produce population-level changes under each intervention, relative to BAU.

### Interventions

#### Main interventions

Table 1 details the impact of the TFG and T21 policies on smoking uptake transition rates. Both interventions were assumed to have the same relative impact on uptake rates by social group. We assumed no change to illicit smoking uptake (from no product use) among youth under either policy. This assumption was tested in sensitivity analyses, where a) each policy was assumed to also reduce illicit market smoking uptake (with half the absolute effect size applied to smoking uptake from the legal market), or b) each policy was instead assumed to increase illicit market uptake (with half those who are prevented from taking up smoking from the legal market due to the policy instead sourcing tobacco from illegal sellers). The two interventions were each compared to the main BAU scenario, alone or in combination with increase illicit market enforcement, detailed below.

**Table 1.**
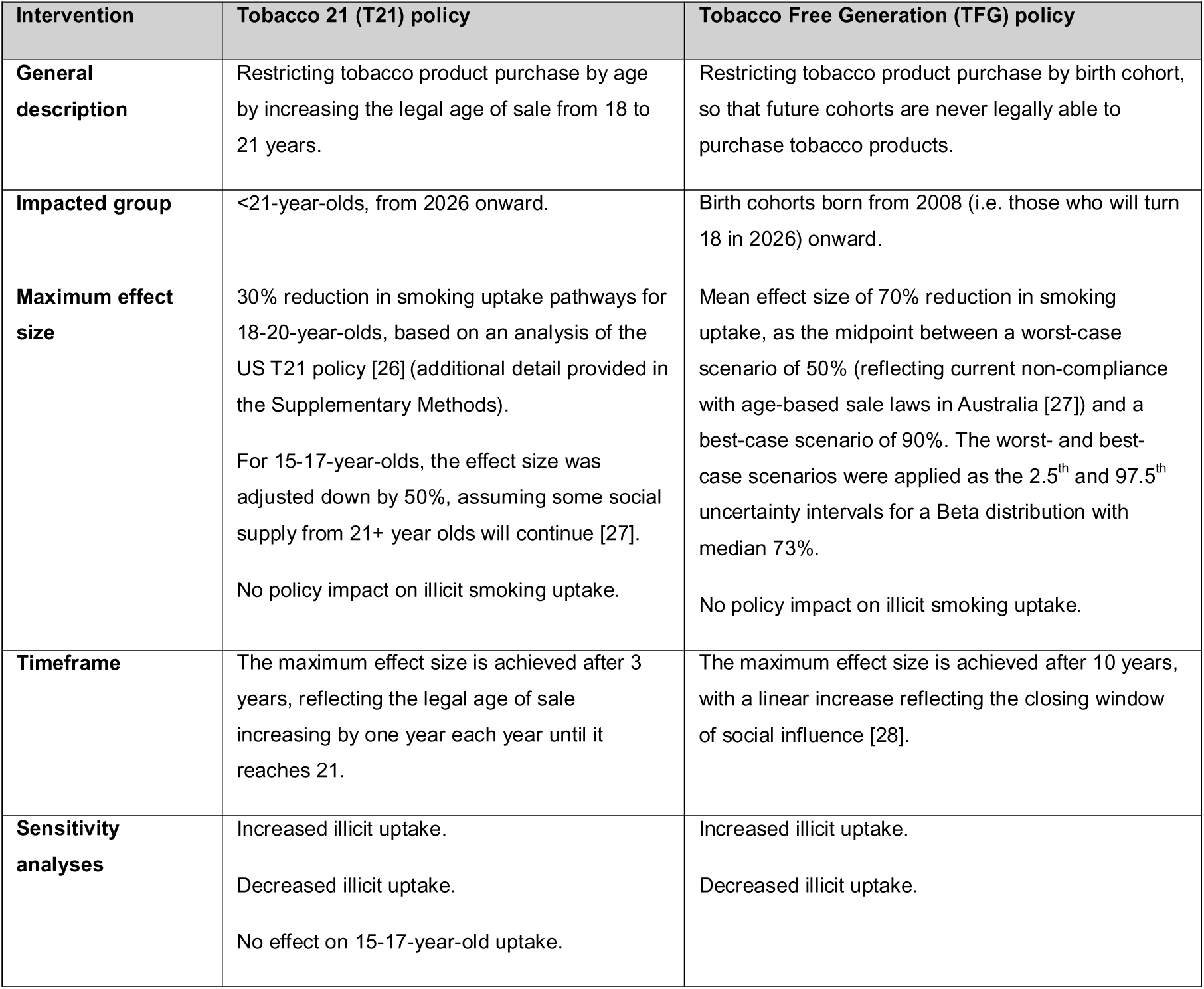
Policy intervention parameterisation.

| Intervention | Tobacco 21 (T21) policy | Tobacco Free Generation (TFG) policy |
| --- | --- | --- |
| **General description** | Restricting tobacco product purchase by age by increasing the legal age of sale from 18 to 21 years. | Restricting tobacco product purchase by birth cohort, so that future cohorts are never legally able to purchase tobacco products. |
| **Impacted group** | <21-year-olds, from 2026 onward. | Birth cohorts born from 2008 (i.e. those who will turn 18 in 2026) onward. |
| **Maximum effect size** | 30% reduction in smoking uptake pathways for 18-20-year-olds, based on an analysis of the US T21 policy [26] (additional detail provided in the Supplementary Methods).<br>For 15-17-year-olds, the effect size was adjusted down by 50%, assuming some social supply from 21+ year olds will continue [27].<br>No policy impact on illicit smoking uptake. | Mean effect size of 70% reduction in smoking uptake, as the midpoint between a worst-case scenario of 50% (reflecting current non-compliance with age-based sale laws in Australia [27]) and a best-case scenario of 90%. The worst- and best-case scenarios were applied as the 2.5<sup>th</sup> and 97.5<sup>th</sup> uncertainty intervals for a Beta distribution with median 73%.<br>No policy impact on illicit smoking uptake. |
| **Timeframe** | The maximum effect size is achieved after 3 years, reflecting the legal age of sale increasing by one year each year until it reaches 21. | The maximum effect size is achieved after 10 years, with a linear increase reflecting the closing window of social influence [28]. |
| **Sensitivity analyses** | Increased illicit uptake.<br>Decreased illicit uptake.<br>No effect on 15-17-year-old uptake. | Increased illicit uptake.<br>Decreased illicit uptake. |

#### Illicit market intervention

Given the uncertainty in future illicit tobacco and e-cigarette market availability/enforcement in Australia, we additionally modelled a scenario of decreased illicit market product supply, assumed to be the result of increased enforcement. This scenario was referred to as an alternative BAU or ‘high enforcement’ and was parameterised as a 90% reduction in access to both illicit tobacco and e-cigarette markets over the next five years. The specific effect sizes for each model transition are provided in the Supplementary Methods and described further in Howe et al [14].

### Running the model

The model was run for 40 years using a Monte Carlo simulation in Python (v3.10.14), drawing from the probability distribution of each uncertain input parameter (Supplementary Table 1). We limited running the model for any longer than this timeframe, as modelled trends become increasingly uncertain with time.

Outcomes produced by the model were changes in deaths and health-adjusted life years (HALYs) under each background enforcement plus intervention combination compared to BAU. HALYs are calculated with the same mathematical form as quality-adjusted life years but use disability weights rather than utility [16]. Age-standardised HALYs were calculated for comparisons across SEIFA quintile, directly to the Australian standard population [29]. Finally, potential effect differences by a) presence of increased illicit market enforcement, and b) SEIFA quintile, on the two main interventions were measured in absolute and relative terms [30]. The main results are undiscounted; HALY gains discounted at 3% per annum are shown in the Supplementary Results.

### Ethics approval

Ethics approval was not necessary for this modelling analysis, as only publicly available data was used (exception: unit-level data from the National Drug Strategy Household Survey, obtained via request from the Australian Data Archive).

## Results

### Policy impacts on smoking and vaping prevalence

Figure 1 presents the impact of the two main interventions on daily smoking prevalence, compared to our base BAU scenario (i.e. with static supply/use of illicit products, reflecting no future increase in illicit market enforcement). Under the TFG policy, daily smoking is expected to decrease to 5.3% (95% uncertainty interval [UI] 5.1-5.50%) in 20 years’ time, 0.7 percentage points (pp) lower than that under the BAU scenario (6.0%). The policy effect is larger for younger ages, with daily smoking prevalence among 15-24-year-olds decreasing to 5.3% (4.8-6.0%) by 2045, compared to 7·6% without the policy.

**Figure 1.**
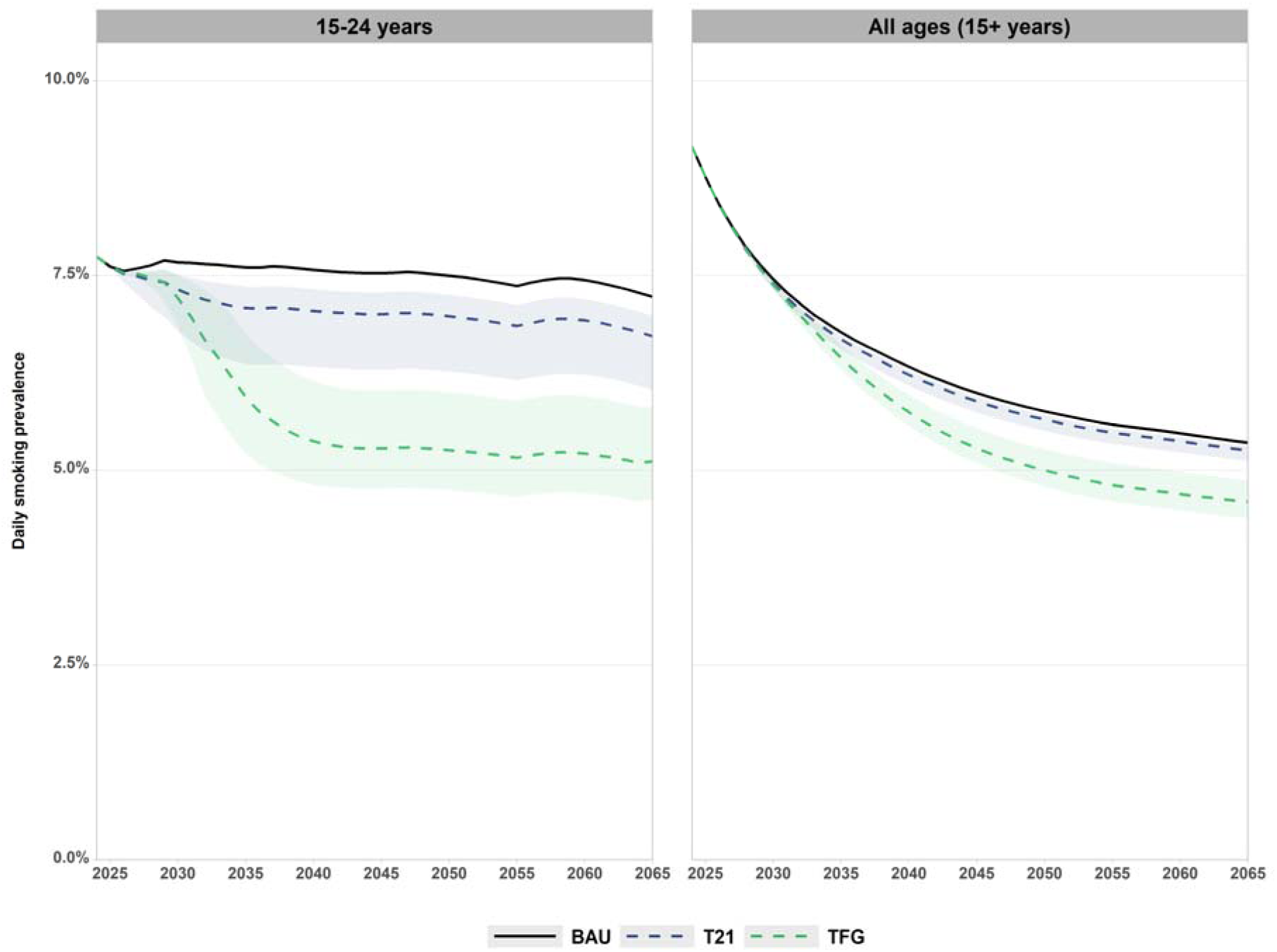
Daily smoking prevalence by age group and policy scenario, 2025-2065. Interventions initiated in 2026. Shaded area reflects the 95% uncertainty interval for the two policies (noting that there is no uncertainty around the BAU projections). Values across all ages shown in Supplementary Figure 2 & Supplementary Table 3. BAU: business-as-usual; TFG: Tobacco free generation.

The T21 policy reduced daily smoking prevalence to 5·9% (5·7-5·9%) in 20 years’ time among 15+ year olds, and to 7% (6·3-7·3%) among 15–24-year-olds specifically. These equated to 0·1 and 0·6pp reductions compared to BAU. Thus, the TFG policy decreased absolute daily smoking prevalence by about 7 times more than the T21 policy for all ages combined, and 3.8 times for 15-24-year-olds specifically.

Daily saw minimal change under either policy, except for a slight increase in vaping occurred among 25-45-year-olds under the TFG policy in the first 20 years (Supplementary Figure 2, Supplementary Table 4).

### Policy impacts on health outcomes

Over 40 years the TFG intervention is expected to result in 75,000 (95% UI 41,900-124,000) health adjusted life years (HALYs, undiscounted) being gained, and 459 (262-663) deaths being averted, when compared to BAU. The TFG policy is expected to result in 6.8 times the HALY gain, and avert 400 additional deaths, compared to the T21 intervention over this period (Table 2, Supplementary Table 6).

**Table 2.** Health-adjusted life years gained by illicit market enforcement level, for T21 and TFG policies.

| BAU scenario | Timeframe | No policy | T21 | TFG |
| --- | --- | --- | --- | --- |
| **Low illicit market enforcement\*** | 2026-2045 | Reference | 2,710<br>(1,140-6,960) | 12,100<br>(6,110-21,600) |
|  | 2046-2065 | Reference | 8,260<br>(3,570-20,600) | 62,900<br>(35,800-102,000) |
|  | Total (40 years) | Reference | 11,000<br>(4,710-27,600) | 75,000<br>(41,900-124,000) |
| **High illicit market enforcement** | 2026-2045 | 137,000<br>(102,000 to 187,000) | 160,000<br>(115,000-220,000) | 165,000<br>(118,000-225,000) |
|  | 2046-2065 | 516,000<br>(388,000 to 689,000) | 670,000<br>(509,000-872,000) | 714,000<br>(540,000-936,000) |
|  | Total (40 years) | 652,000<br>(491,000 to 874,000) | 829,000<br>(627,000-1,090,000) | 879,000<br>(660,000-1,160,000) |

\*Main BAU scenario
BAU: business-as-usual; TFG: Tobacco free generation; T21: Tobacco-21.

Increasing the relative harm of vaping to 40% of smoking-related disease risk resulted in an approximately 19% decrease in health gains from the TFG intervention (Supplementary Table 7), resulting from the increase in vaping under this intervention (Supplementary Figure 2).

### Sensitivity analyses

In the results described above, both interventions were parameterised assuming no change to uptake of smoking tobacco from illicit sources among the affected cohorts. In Supplementary Figures 3-4, the impact on smoking prevalence is shown under two variations to this assumption.

In the first scenario, illicit smoking initiation *increased* under both policies. For the TFG policy, this results in daily smoking reducing to 6·8% for 15-24-year-olds, 1.5pp less than the reduction simulated in the main policy scenario. Associated health gain in HALYs was 64% lower than that in the main policy scenario (Supplementary Table 9). For the T21 policy, daily smoking reached 7·3% by 2045 among 15-24-year-olds, 0·3pp less than the effect of the main policy scenario. HALY gain was 61% lower than the main scenario.

In the second scenario, illicit smoking initiation *decreased* under both policies. For the TFG policy, this could see daily smoking reducing to 4.3% by 2045 for 15-24-year-olds, an additional 1.0pp decrease on top of the effect seen with no change to illicit supply. This increased HALY gain by 33% on top of that seen in the main policy scenario (Supplementary Table 9). For the T21 policy, daily smoking reaches 6·7% by 2045 for 15-24-year-olds, a 0·3pp additional reduction than the main analysis. HALY gain was 60% higher than in the main policy scenario.

In an additional sensitivity analysis, with no impact of the T21 intervention on uptake among <18-year-olds, daily smoking among 15-24-year-olds was modelled to reach 7.2% by 2045 (Supplementary Figure 4, Supplementary Table 8). HALY gain reduced by 30% relative to the main policy scenario (Supplementary Table 9).

Finally, the two policies were modelled on top of alternative BAU parameterisation options. Despite wide differences in these BAU trends, the absolute and relative reduction in smoking prevalence under each intervention remained consistent with the main analysis (Supplementary Figure 5, Supplementary Table 10).

### Policy impacts by illicit market enforcement level

Under the combined TFG policy with high illicit market enforcement (modelled as a 90% reduction in illicit market product use), smoking prevalence among 15-24-year-olds was estimated to reduce to 1.3% (95% UI 0.8-2.0%) by 2045, compared to 3.5% with high illicit market enforcement alone and 5.3% with the TFG policy alone (Figure 2). This means that if high enforcement of illicit markets is in place, a TFG policy may reduce daily smoking prevalence in this age group by 2.1 times what is achievable from a TFG policy in a setting of low enforcement (63% reduction from TFG under high enforcement scenario vs. 30% reduction from TFG under low enforcement; Supplementary Table 12).

**Figure 2.**
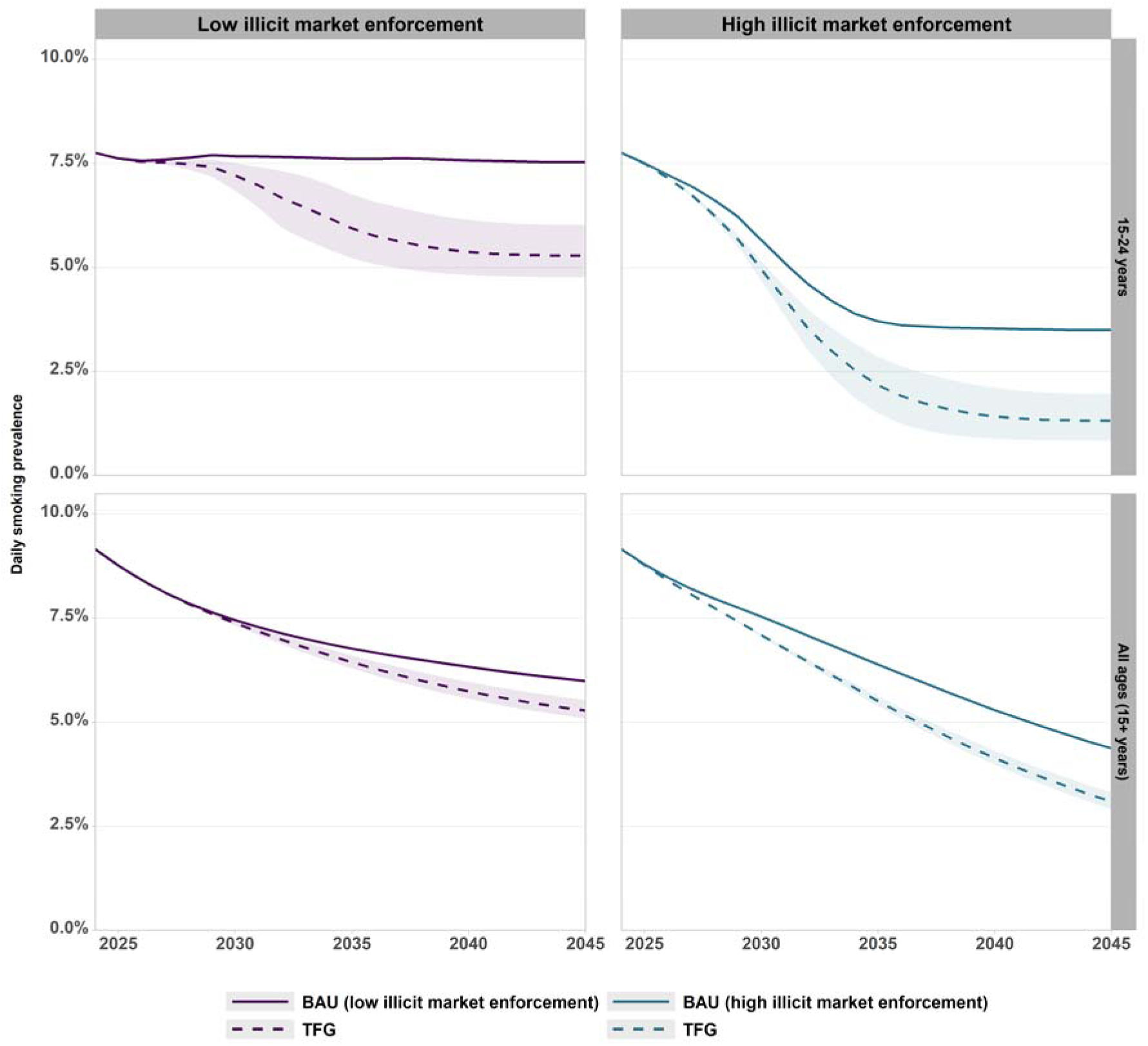
Daily smoking prevalence by age group, comparing TFG under varied background illicit market enforcement, 2025-2045. TFG initiated in 2026. Shaded area reflects the 95% uncertainty interval for the policy (noting that there is no uncertainty around the BAU projections). Values shown in Supplementary Table 11. BAU: business-as-usual; TFG: Tobacco free generation.

Under the combination of high illicit market enforcement and the TFG policy, daily vaping prevalence after 20 years was 0.7pp greater than under high illicit market enforcement alone (Supplementary Table 13). A similar effect was seen under the combined T21 + high enforcement scenario. This happened as a downstream result of decreased switching from vaping to illicit tobacco that occurred under high enforcement; a greater number of young people in the vaping state resulted in a larger impact from the two policies reducing movement from vaping to legal smoking. However, vaping in this scenario was still lower than under that projected under the low enforcement BAU (Supplementary Figures 6 & 8).

Health gains under the variation in illicit market enforcement are shown in Table 2.

### Policy impacts by SEIFA quintile

Over time under BAU, the absolute gap in daily smoking prevalence (all 15+ year olds) between the most disadvantaged and most advantaged SEIFA quintiles is expected to decrease, from 11·4pp in 2025 to 7·6pp in 2045 (Figure 3). The TFG scenario reduced the gap in 2045 further to 7.0pp. For 15–24-year-olds, the gap was reduced by 27% by 2045, to 6.4pp compared to 8·8pp under BAU. This reflects a 2.4pp greater impact of the TFG policy for the SES1 compared to SES5 quintile (Supplementary Table 18). Health gains across SEIFA quintiles are shown in Supplementary Table 20.

**Figure 3.**
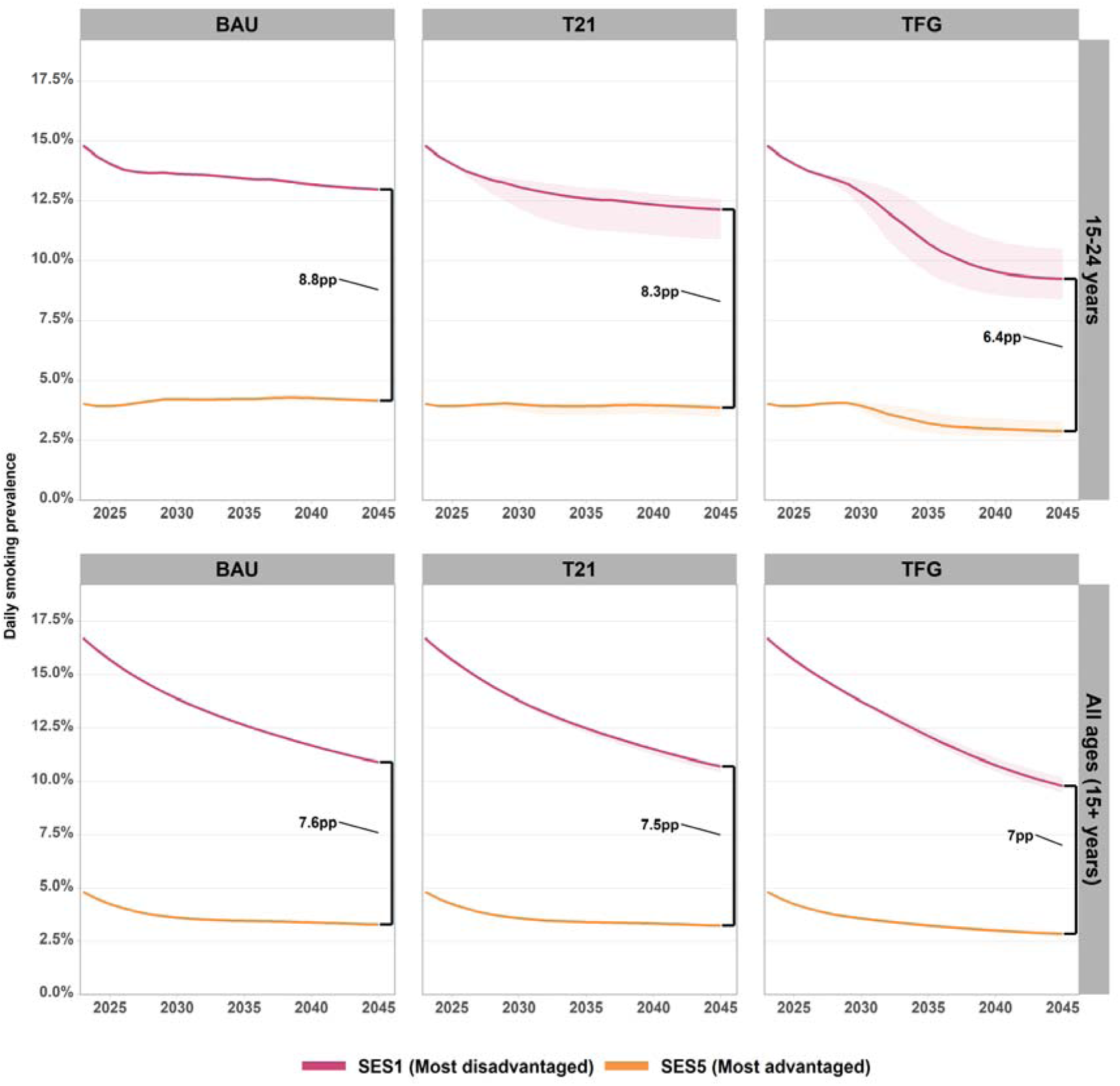
Daily smoking prevalence by age group and policy scenario, comparing SEIFA quintiles. Interventions initiated in 2026. Difference between SES1 and SES5 in 2045 across scenarios shown in percentage point (pp) terms. Shaded area reflects the 95% uncertainty interval for the two policies (noting that there is no uncertainty around the BAU projections). Values across all ages shown in Supplementary Figure 9 & Supplementary Table 17. BAU: business-as-usual; TFG: Tobacco free generation.

## Discussion

This modelling study found that a birthdate-based tobacco sale restriction, or tobacco free generation (TFG) law, is likely to be more effective than increasing the minimum legal sale age from 18 to 21 years, in terms of future smoking rates and population health impacts.

However, the effect of the TFG policy was sensitive to the background level of illicit tobacco uptake. In a setting of high enforcement of illicit tobacco and e-cigarette supply (modelled as a reduction in availability of these products by 90%), both modelled policies produced larger relative reductions in smoking prevalence and larger absolute health gain than that achievable by either policy in a setting of continued wide illicit market product availability. Further, under a pessimistic scenario in which half of those who would have taken up smoking from the legal market instead doing so via illegal sellers under a TFG law, the impact on smoking prevalence reduced by 65% among 15-24-year-olds. This highlights the importance of ongoing actions to reduce illicit market supply to support other tobacco control policies.

Assuming the relative impacts of interventions by socioeconomic status are the same, both TFG and T21 policies were projected to have pro-equity effects, reducing the absolute gap in smoking prevalence between socioeconomic groups and generating larger health gains among more disadvantaged groups for both absolute and relative health gains. This inequity reduction remained under the increased background illicit enforcement scenario.

Our results are broadly consistent with international modelling studies showing that age- or cohort-based tobacco sales restrictions can reduce future smoking prevalence and tobacco-related disease, but direct comparison is difficult because intervention assumptions differ substantially. Kim et al. assumed 100% compliance with a TFG law in South Korea with no further smoking uptake for included cohorts [9]. In contrast, Davies et al. estimated a much more conservative effect size at 23% reduction in uptake for 18+ year olds under a smokefree generation policy in England, based on T21 policy effect estimates [31]. Modelling of the Aotearoa/New Zealand endgame package, using an earlier version of the SHINE-Tobacco model, applied a 90% reduction in uptake, assuming a limited amount of social supply would continue [12]. Although absolute impacts vary, predicted pro-equity effects of a TFG law were similarly seen in other research, across Indigenous status in New Zealand [12], and social deprivation area in England [31]. Future evidence from jurisdictions that have recently implemented TFG laws will generate real-world evidence to improve simulation modelling.

Our findings support the potential role of TFG as part of an Australian tobacco endgame strategy, but they should not be interpreted as implying that the primary rationale for TFG is short-term health gain. TFG is a cohort-based endgame policy, with its justification including prevention of addiction before it starts and denormalisation of tobacco supply, in addition to projected health gain. This distinction is important given that most of the morbidity and mortality reductions occur decades after the policy is introduced, after the included cohorts age into periods of higher tobacco-related disease risk.

Further, a TFG law is just one part of the solution to the tobacco epidemic. Even if youth do not take up tobacco use, the remaining number of people who currently smoke is substantial, providing a continuing market for the tobacco industry and providing the conditions that support ongoing illegal supply from older cohorts and illegal sales to youth. Interventions that increase smoking cessation among all ages are required for more immediate reductions in smoking prevalence [18], and to reduce the viability of the tobacco market (both legal and illicit) in Australia.

### Limitations

The study also has important limitations. First, there is currently little real-world evidence on the population-level impact of TFG laws, so intervention effect sizes were necessarily hypothetical and based on plausible assumptions rather than direct evidence. Second, the uncertainty intervals reported reflect uncertainty in selected input parameters, but not structural uncertainty. Further, the BAU Markov model transitions did not include uncertainty due to a lack of available input data, with some behavioural transition parameters being informed by international data. As a sensitivity analysis, the T21 and TFG interventions were compared against variations in the BAU parameterisation. Despite large changes in the BAU trends under these different options, the relative and absolute impact of the interventions remained consistent with the main analysis.

Third, long-term disease risks from vaping remain uncertain; our assumptions about relative disease risk are consistent with previous modelling, but future epidemiological evidence may alter these estimates. Fourth, the illicit market enforcement scenarios are hypothetical. They should be interpreted as scenario analyses showing how reduced illicit supply could modify policy impact, rather than predictions of the effect of any specific enforcement package.

Finally, we applied the T21 and TFG effect sizes uniformly across SEIFA quintiles. However, impacts may vary between SEIFA quintiles, for example, if social supply is higher in low-income areas, or if enforcement of the policy is not as strong. If this is the case, the pro-equity effect estimated in this analysis would be an overestimate. Additional research is required to test possible differences in policy compliance across SES, as well as other dimensions of equity, including First Nations status and geographic remoteness, which are highly relevant to Australian tobacco control and can be examined in future with the SHINE-Tobacco model.

## Conclusion

A TFG policy could make an important long-term contribution to reducing smoking prevalence, tobacco-related disease and socioeconomic inequities in Australia. Its impact would be greatest if introduced alongside effective measures to reduce illicit tobacco and e-cigarette supply and policies that increase cessation among people who already smoke. TFG should therefore be viewed neither as a standalone solution nor as a short-term health-maximising intervention, but as a prevention-focused endgame policy whose benefits depend on the broader tobacco and nicotine market in which it is implemented.

## Data Availability

This article includes no original data. Requests to the corresponding author for the model code will be considered on a case-by-case basis.

