## Supplementary Material for "Modelling a tobacco-free generation policy in Australia: population health impacts under illicit market uncertainty"

### Supplementary Methods

#### 1. SHINE-Tobacco model overview

SHINE-Tobacco consists of a discrete time Markov model to describe smoking and vaping behaviours in yearly time steps, and a proportional multi-state lifetable (PMSLT) used to quantify the health impacts of changes to smoking-related disease rates.

Business-as-usual scenarios (BAU) are first specified for both model components – this involves projecting forward tobacco-related disease rates, all-cause mortality rates, and population counts (the PMSLT component) and calibrating transitions to and from smoking and vaping behaviours (the Markov model component) under the assumption that the momentum from tobacco control policies applied over the past 20 years will continue into the future. Intervention scenarios can then be modelled on top of the BAU scenario – this occurs through making changes to smoking or vaping transitions (e.g., reducing the smoking uptake rate) in the Markov model, which flows through to changes in tobacco-related disease incidence in the PMSLT, and resultingly changes to population health over time.

The previous iteration of this model applied in the Australian context (1) only included smoking. The present version has been updated to incorporate two additional factors:

1. **E-cigarette use.** Vaping rates have risen in Australia from 2016-2023, particularly for youth. (2, 3) While regulatory changes introduced in late 2024 have been made to restrict e-cigarette availability to pharmacies, it is unclear how effective these will be at reducing illicit e-cigarette use. Most recent estimates (prior to the policy change) had the majority of e-cigarette supply occurring via the illicit market. (4)
2. **Illicit tobacco market use.** We define illicit tobacco products as those that are untaxed and/or without plain packaging. This does not include the sale of taxed cigarettes to underage customers (minimum age of purchasing tobacco is currently 18 years in Australia). The proportion of tobacco consumption that is via illicit supply has increased considerably in recent years – the most recent estimate from the Australian Taxation Office (ATO) was 25% of total consumption in 2023-24. (5) A more recent report from the federal government’s Illicit Tobacco and E-cigarette Commissioner estimated illicit tobacco use as somewhere between 50-60% of total consumption in 2024-25. (6)

Given the uncertainty surrounding both factors, we parameterise multiple BAU scenarios reflecting possible changes to the enforcement of illicit e-cigarettes and tobacco. These scenarios were explored in detail in a separate publication (currently a pre-print). (7) For the present analysis, we use the different scenarios as alternative comparison scenario, providing varying background levels of illicit market enforcement on top of which a tobacco control policy may be applied.

#### 2. BAU model inputs

Model inputs are detailed in Supplementary Table 1. Application of each input in the model is described in further detail in the following sections.

**Supplementary Table 1. Model input parameters.**

| **Input** | **Parameterisation** | **Uncertainty** |
| --- | --- | --- |
| Smoking Markov model transitions | Yearly net uptake and cessation rates were fit to daily smoking prevalence forecasts (produced based on synthetic dataset produced in Howe et al. (8). Note that input data was limited to a defined ‘pre-vaping’ period (2001-2016) using a simulated annealing optimisation model.  Rates are held constant after 20 years. | No uncertainty in main smoking transitions |
| Vaping & dual use Markov model transitions | Base year uptake, cessation, and switch rates were fit to recent daily vaping prevalence (produced based on logistic regression applied to 2016-2022/23 data by sex and age) using a simulated annealing optimisation model.  Calibrated rates disaggregated by socioeconomic strata based on VSHS data (9), then projected forward to reflect the ‘low enforcement’ scenario. E-cigarette transitions were then adjusted to reflect the ‘high enforcement’ scenario (7). | Two scenarios of varied vaping and dual use transitions modelled as possible variations to illicit supply  Two additional scenarios varying smoking-vaping switch rates included as sensitivity analyses |
| Illicit tobacco market transitions | Yearly smoking uptake and cessation rates disaggregated into legal vs. illicit states based on the price elasticity of tobacco (10)* and relative price difference of legal and illicit products. Switching from legal to illicit sources of tobacco was then calibrated so that the proportion of overall smoking occurring via illicit supply increased from 25% in the base year to 50% in 2025, reflecting recent trends.(5,6) This reflects the ‘low enforcement’ scenario.  Transitions into/out of the illicit states were then adjusted to reflect the ‘high enforcement’ scenario (7). | Two scenarios of varied illicit smoking use transitions modelled as possible variations to illicit supply |
| Start year Markov model prevalence values | Total smoking by social group estimated from regression on 2001-2022/23 synthetic smoking dataset (8).  Naïve vaping, non-naïve vaping, and dual use by sex and age based on regression on NDSHS data from 2016-2022/23; disaggregated into social groups based on VSHS data (9) (assuming uniform difference by age).  Smoking and dual use prevalence were split into illicit and legal sub-states based on 25% of total smoking consumption being sourced via the illicit market in 2023 (5) (adjusted down to account for some of this being through higher smoking intensity in illicit states). | No uncertainty |
| Smoking intensity | Regression model applied to unit-level NDSHS data (obtained from Australian Data Archive (11)) to estimate cigarettes smoked per day by birth cohort and social group (as per Howe et al. (1, 12)).  Vaues adjusted for illicit vs. legal markets based on price elasticity*. Pack-years then calculated for each birth cohort, with age of uptake set to 20. | 95% uncertainty interval based on bootstrapping of the regression model (1, 12) |
| Vaping intensity | Derived from base year cigarettes per day. Assumed to be static over time. For vaping ‘pack-year’ calculation, assume e-cigarettes smoked per day is zero prior to 2017. | Same as smoking intensity |
| Smoking-disease RRs | Values for 31 tobacco-related disease, by cigarettes smoked per day or pack-year risk units, from Dai et al. (13) (GBD data, downloaded from IHME website (14)). | 95% uncertainty interval based on confidence interval from GBD data |
| Vaping-disease RRs | Derived from smoking-disease RRs, scaled down to 11% of the mean smoking RR (15). | 95% uncertainty interval defined by scaling smoking RRs down to 5% and 20%. |
| Disease rates | Incidence, case fatality, and remission rates projected forward based on historic (1990-2021) GBD data (16), using method by Wilson et al. (17)  Disease disability weights derived from GBD 2021 (16) (as years lived with disability/cases); held constant in future years. | 95% uncertainty interval based on confidence interval from GBD data |
| All-cause morbidity and mortality | Mortality projected forward based on historic (1990-2021) GBD data. (16) Values held constant after 20 years.  Population-level morbidity derived from 2021 GBD (16) data (as years lived with disability summed across all causes/population count); held constant in future years. | 95% uncertainty interval based on confidence interval from GBD data |

*Price elasticity set at -0.4 (10), split evenly between change in smoking intensity and use. For a price reduction of 65% (based on average costs of $43 (18) for legal cigarettes vs. estimate of $15 for illicit (19)), 21% change in intensity (increase) and quit rate (decrease) for illicit compared to legal smoking.

#### 3. Markov model

The SHINE-Tobacco Markov model consists of yearly transition rates between different smoking and vaping behaviour states. The different transitions that can occur are shown in Supplementary Figure 1. For each product use state, we specifically model only daily use, as a conservative estimate of subsequent health impacts. Not shown in this figure are the transitions that occur from each state to a death state. For transitions from smoking to non-smoking states (either quitting, or switching to vaping), 30 tunnel states are used to overcome the memoryless assumption of Markov models, with a gradual reduction in tobacco-related disease risk being set in each of the tunnel state as years since quitting increases. (20)

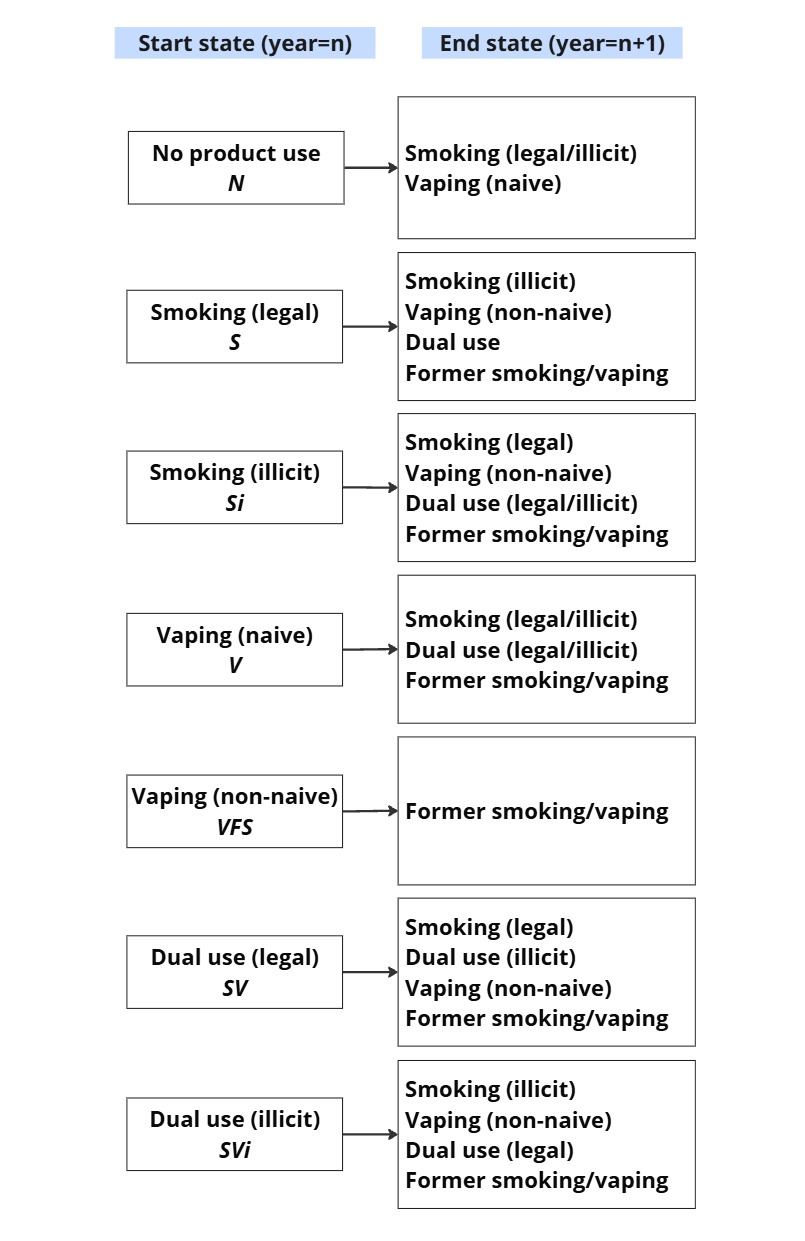

**Supplementary Figure 1. Markov model transitions.**

The smoking states are split into ‘legal’ and ‘illicit’, to differentiate between tobacco products sold legally vs. through illicit-market supply. Illicit tobacco products are those that have not paid customs duties and do not follow product standards required for sale in Australia (e.g., do not have plain packaging).

Vaping states are split into ‘naïve’ and ‘non-naïve’ states, to differentiate between vaping uptake by those who have never previously smoked daily, compared to vaping uptake as an alternative to smoking. Note that the naïve state is inherently an illicit state, as e-cigarettes are not legally available in retail settings outside of controlled product provision in pharmacies for smoking cessation. The non-naïve state incorporates some legal supply via pharmacies, but predominantly e-cigarettes sourced via the illicit market.

The Markov model is parameterised for a ‘business-as-usual’ (BAU) or base case scenario in a three-step process:

1. Calibrate total smoking uptake and cessation rates, assuming that rates will follow the projected downward trend in smoking prevalence. (8)
2. Calibrate vaping uptake and cessation rates, and switch rates between smoking, vaping and dual use, assuming that rates will remain constant based on current estimates of vaping prevalence.
3. Separate the calibrated smoking transitions and smoking intensity into illicit vs. legal states based on the price difference in illicit vs. legal products. Switching from legal to illicit sources of tobacco was then calibrated so that the proportion of overall smoking occurring via illicit supply increased from 25% in 2023 to 50% in 2025, reflecting recent trends. (5,6)

As a sensitivity analysis, two alternative BAU parameterisations were applied, involving changing the strength of relationship between switching from smoking to vaping, or vaping to smoking (as per (7)). Additional detail on calibration of the Markov model is available in previous work ((12) and (7) [pre-print]).

#### 4. PMSLT

The PMSLT used for this model is well-established and is described in detailed in Blakely et al. (21) Its application for the SHINE-Tobacco model is also detailed in Howe et al. (1) The purpose of the PMSLT is to calculate the disease-specific and/or all-cause health impacts that occur for a population over time under an intervention scenario compared to status quo or ‘business-as-usual’ (BAU). It is composed of a main all-cause lifetable, and subsidiary disease lifetables. (21)

The population is modelled in the main lifetable, by strata of sex, age, remoteness, SEIFA, and Indigenous status, for 40 years (with the main results focusing on the first 20 years). The population count and composition in each year under BAU is determined by yearly births, migration, and projections of all-cause mortality by strata (described in (1)). Population-level morbidity is determined by time-invariant disability rates. Alongside the main lifetable are 31 smoking-disease lifetables, comprised of projections of incidence, case fatality, and remission rates, alongside time-invariant disability rates, by strata. Each rate input is forecast for 20 years, then held constant for the subsequent 20 years.

Each stratum in the model is further disaggregated by smoking/vaping status, determined by the Markov model as described in the previous section. Each Markov model cohort carries a different risk of each smoking-related disease. These smoking-disease relationships are input as incidence rate ratios (RRs) that follow dose-response trends depending on smoking intensity and duration. Under an intervention scenario, changes to smoking prevalence in the Markov model are combined with the RRs in a population-impact fraction (PIF). The PIF results in a change in disease incidence, which flows through to changes in disease-specific, then all-cause, morbidity and mortality.

The PIF is calculated in each year and for each stratum as

$$PIF_{sdt} = \frac{\sum_{j=1}^{n} P_{sjt} RR_{sdj} -\sum_{j=1}^{n} P_{jt}^{'} RR_{sdj}}{\sum_{j=1}^{n} P_{sjt} RR_{sdj}}$$

where $s$ is the stratum (sex by age by remoteness by SEIFA by Indigenous status), $t$ is the time step (year), $d$ is the disease, $P$ is the proportion of the specific strata in each state model state $j$ (out of $n$ states) under BAU compared to the intervention scenario ($P’$), and $RR$ is the relative risk linking smoking rates by strata to the incidence of each disease.

The dose-response curves, sourced from the GBD (14), for smoking-disease incidence relies on pack-years (derived from cigarettes smoked per day and number of years smoking) for cancers and chronic obstructive pulmonary disease (COPD) cigarettes, and on cigarettes smoked per day directly for other diseases. For the SHINE-Tobacco model, cigarettes per day has been projected both forward and backward, in order to calculate ‘pack-years’ for each cohort in the model (e.g., for those aged 80 in 2025, if we assume smoking uptake occurred at 20 years of age, we need to back-project cigarettes smoked per day for 1965-2025 for current pack-year estimation). A logistic regression model was applied to National Drug Strategy Household Survey (NDSHS) data from 2001 (earliest unit-level data available)-2022/23 for this purpose. The method is described in detail in previous work (1, 12).

For this analysis, smoking is further disaggregated into legal vs. illicit sub-states. Total cigarettes per day estimates were adjusted to reflect higher smoking intensity in the illicit states (based on the difference in price) then used to calculate pack-years for each stratum and state.

Vaping intensity, in terms of ‘e-cigarettes per day’ was assumed to be equivalent to cigarettes per day estimated in 2022 for a given sociodemographic group. Unlike for smoking, we assumed that vaping intensity is static (i.e., no change in e-cigarettes smoked per day in the future). To estimate e-cigarette ‘pack-years’ for a cohort, the number of years the cohort has been vaping is required. Given that vaping is a more recent trend than smoking (in the model we assume that no vaping occurred prior to 2016), for any cohort that turned 20 prior to the model start year, uptake was assumed to have started in 2020 (the approximate midpoint year that uptake could have occurred). For any future cohort, pack-years for e-cigarette use is calculated assuming that uptake occurred at age 20.

#### 5. Illicit market enforcement scenario

The illicit market enforcement scenario was defined in our previous work (currently a pre-print). (7) The scenario, assumed to reflect high enforcement of the illicit market, was specified as per Supplementary Table 2.

**Supplementary Table 2. Illicit market enforcement specification.**

| **Transition** | **Impact of high enforcement intervention** |
| --- | --- |
| **Illicit e-cigarette enforcement** | |
| Naïve vaping uptake rate | RR = 0.1  90% reduction assumed to be maximum possible enforcement level |
| Non-naïve vaping uptake rate (i.e., switching from smoking) | RR = 0.23  77% reduction (scaled down from above to account for approximately 14% of non-naïve vaping occurring via the legal pathway (2)) |
| Naïve vaping cessation rate | RR = 3.16  Half the absolute impact (log scale) on vaping uptake, assuming that it is harder to quit vaping than not start |
| Non-naïve vaping cessation rate | RR = 2.86 (scaled down from above to account for approximately 14% of non-naïve vaping occurring via the legal pathway (2)) |
| Switching from naïve vaping to smoking | RR=1.93  This is 0.57 times the impact on quitting vaping, per finding from Posner et al. (22) |
| Switching from dual use to smoking | RR = 8.74  Same as the absolute impact (log scale) of switching from smoking to vaping (i.e. non-naïve vaping uptake) |
| **Illicit tobacco enforcement** | |
| Illicit smoking uptake | RR = 0.1  90% reduction assumed to be maximum possible enforcement level |
| Illicit smoking cessation | RR=1.19  Based on price elasticity (estimated as a decrease in use by 20.7% under price change from illicit to legal), applied to 90% of those smoking illicit tobacco. |
| Switching from illicit smoking to legal smoking | 90% of those remaining in the illicit smoking state, after the above cessation impact, are moved into the legal smoking state. |
| Smoking intensity in illicit smoking states | Reduced by 20.7% (i.e., the price elasticity-based multiplier), for 90% those who remain in the illicit smoking states. |

Values reached after 5 years (gradual increase starting in 2025).

RR = relative risk.

Based on the 2022-23 National Drug Strategy Household Survey, 14% of 18+ year olds who vaped daily used a prescription to source e-cigarettes. (2) We incorporated these different sources in the modelled high enforcement scenario by a) scaling down effect sizes for transitions into/out of the non-naïve vaping state to account for this proportion of individuals who are using the pharmacy pathway (fixed at 14%) who will not be affected by increased enforcement.

The illicit tobacco enforcement estimates are based on a price elasticity of tobacco of -0.4 (10). This elasticity is assumed to be split evenly between smoking intensity, and tobacco use (yes/no), i.e., a price elasticity of -0.2 for each component. At a price difference of $0.75 (19) vs. $2.11 (23) per stick for illicit vs. legal markets, or a 64% price reduction, we translated this to a $-0.2*(\ln\left( 1-0.64 \right))=$20.7% reduction in smoking intensity, and 20.7% increase in cessation (Supplementary Table 2).

#### 6. T21 policy analysis

We aimed to identify the impact of age-based laws on smoking participation (uptake and/or quit rates, or prevalence). We included the impact for both 18-20-year-olds directly affected by the policy, and other ages (<18-year-olds, and 21+ year olds). We excluded studies looking at specific sub-populations (e.g. smoking in pregnancy).

A 2024 systematic review was identified as assessing the impact of minimum legal sale age laws on smoking (24), though this did not include a meta-analysis. The papers within this study were reviewed, as well as additional studies identified published after the systematic review (up to 01/04/2026, using the search terms from (24)). The additional search identified another systematic review, specifically of the US T21 policy, including meta-analysis; (25, 26) however, there was significant heterogeneity between studies, including specific outcome assessed, follow up times, and policy coverage (with many studies examining local level T21 laws as opposed to state or federal).

The most appropriate analysis identified was from Hansen et al. (27), assessing the impact of statewide adoption of the US T21 policy on smoking prevalence, a minimum of 1-year post-policy introduction. Smoking data from the cross-sectional Behavioural Risk Factor Surveillance System (BRFSS) survey was used in a stacked difference-in-difference (DID) analysis to estimate the impact of the T21 policy on tobacco use. The stacked DID analysis accounts for the staggered adoption of the T21 policy over time across states. In this analysis, daily smoking was found to reduce by 30% among 18-20-year-olds. (27) We assume that the full effect was from reduced uptake in this group (as opposed to increased cessation).

### Supplementary Results

#### 1. Smoking and vaping prevalence

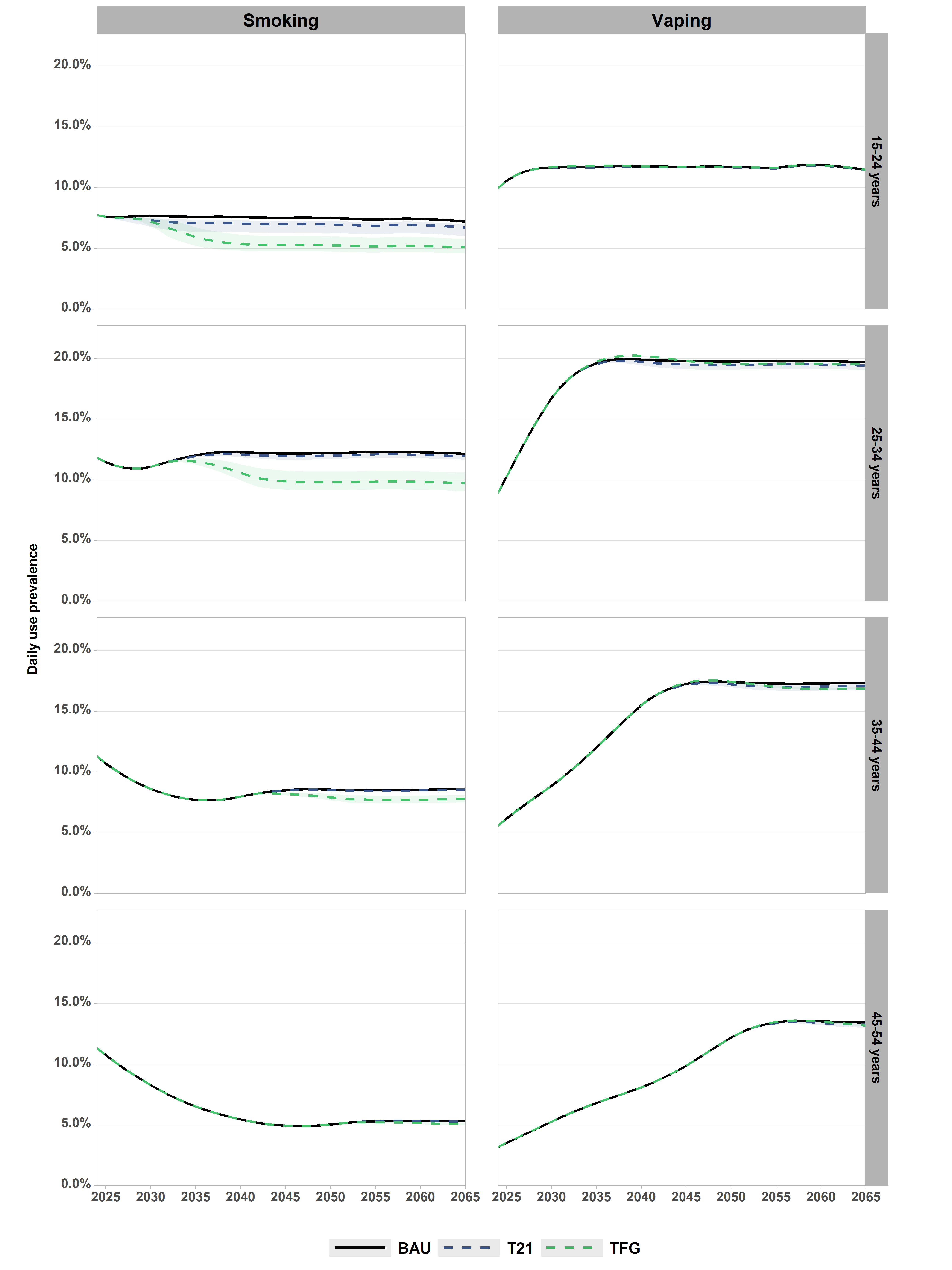

**Supplementary Figure 2. Daily smoking and vaping prevalence by age group, 2026-2065.** Note: age 55+ year olds not shown (no effect of T21 or TFG policy on these age groups in any year). Values shown in Supplementary Table 3. BAU: business-as-usual; TFG: Tobacco free generation; T21: Tobacco-21.

**Supplementary Table 3. Daily smoking prevalence under each scenario, by age and year.**

| **Age group** | **2025*^#^** | **2045** | **2065** |
| --- | --- | --- | --- |
| **BAU (Low illicit market enforcement)^#^** | | | |
| **15+** | 8.80% | 6% | 5.4% |
| **15-24** | 7.60% | 7.60% | 7.5% |
| **25-34** | 11.40% | 11.50% | 12.2% |
| **35-44** | 10.70% | 10.70% | 8.5% |
| **45-54** | 10.70% | 10.70% | 4.9% |
| **55-64** | 8.90% | 9% | 3.5% |
| **65+** | 4.40% | 4.50% | 2% |
| **T21** | | | |
| **15+** | 8.80% | 5.9% (5.7%-5.9%) | 5.3% (5.1%-5.3%) |
| **15-24** | 7.60% | 7% (6.3%-7.3%) | 6.7% (6%-7%) |
| **25-34** | 11.50% | 12% (11.7%-12.1%) | 12% (11.7%-12.1%) |
| **35-44** | 10.70% | 8.5% (8.5%-8.5%) | 8.6% (8.5%-8.6%) |
| **45-54** | 10.70% | 4.9% (4.9%-4.9%) | 5.3% (5.3%-5.3%) |
| **55-64** | 9% | 3.5% (3.5%-3.5%) | 2.9% (2.9%-2.9%) |
| **65+** | 4.50% | 2% (2%-2%) | 1% (1%-1%) |
| **TFG** | | | |
| **15+** | 8.80% | 5.3% (5.1%-5.5%) | 4.6% (4.4%-4.9%) |
| **15-24** | 7.60% | 5.3% (4.8%-6%) | 5.1% (4.6%-5.8%) |
| **25-34** | 11.50% | 9.9% (9.2%-10.8%) | 9.7% (9.1%-10.6%) |
| **35-44** | 10.70% | 8.2% (8.1%-8.3%) | 7.8% (7.5%-8.1%) |
| **45-54** | 10.70% | 4.9% (4.9%-4.9%) | 5.1% (5%-5.2%) |
| **55-64** | 9% | 3.5% (3.5%-3.5%) | 2.9% (2.8%-2.9%) |
| **65+** | 4.50% | 2% (2%-2%) | 1% (1%-1%) |

*Pre-intervention: all values same.

^#^No uncertainty under BAU/pre-intervention

BAU: business-as-usual; TFG: Tobacco free generation; T21: Tobacco-21.

**Supplementary Table 4. Daily vaping prevalence under each scenario, by age and year.**

| **Age group** | **2025*^#^** | **2045** | **2065** |
| --- | --- | --- | --- |
| **BAU (Low illicit market enforcement) ^#^** | | | |
| **15+** | 5.40% | 10% | 11.2% |
| **15-24** | 10.60% | 11.7% | 11.5% |
| **25-34** | 10.30% | 19.8% | 19.7% |
| **35-44** | 6.20% | 17.3% | 17.4% |
| **45-54** | 3.50% | 9.9% | 13.4% |
| **55-64** | 1.90% | 5.1% | 9.5% |
| **65+** | 0.60% | 1.8% | 3.7% |
| **T21** | | | |
| **15+** | 5.40% | 10% (9.9%-10%) | 11% (10.9%-11.1%) |
| **15-24** | 10.60% | 11.7% (11.6%-11.7%) | 11.4% (11.4%-11.5%) |
| **25-34** | 10.30% | 19.5% (19.1%-19.6%) | 19.4% (19.1%-19.6%) |
| **35-44** | 6.20% | 17.2% (17.1%-17.2%) | 17.1% (16.8%-17.2%) |
| **45-54** | 3.50% | 9.9% (9.9%-9.9%) | 13.3% (13%-13.4%) |
| **55-64** | 1.90% | 5.1% (5.1%-5.1%) | 9.4% (9.4%-9.4%) |
| **65+** | 0.60% | 1.8% (1.8%-1.8%) | 3.7% (3.7%-3.7%) |
| **TFG** | | | |
| **15+** | 5.40% | 10% (10%-10%) | 11% (11%-11.1%) |
| **15-24** | 10.60% | 11.7% (11.7%-11.7%) | 11.5% (11.5%-11.5%) |
| **25-34** | 10.30% | 19.8% (19.7%-19.8%) | 19.5% (19.5%-19.5%) |
| **35-44** | 6.20% | 17.4% (17.3%-17.4%) | 16.9% (16.8%-17%) |
| **45-54** | 3.50% | 9.9% (9.9%-9.9%) | 13.2% (13.1%-13.3%) |
| **55-64** | 1.90% | 5.1% (5.1%-5.1%) | 9.5% (9.5%-9.5%) |
| **65+** | 0.60% | 1.8% (1.8%-1.8%) | 3.7% (3.7%-3.7%) |

*Pre-intervention: all values same.

^#^No uncertainty under BAU/pre-intervention

BAU: business-as-usual; TFG: Tobacco free generation; T21: Tobacco-21.

#### 2. Health impact

**Supplementary Table 5. Health-adjusted life years gained under each intervention compared to BAU, by timeframe; discounted at 3% per annum.**

| **Timeframe** | **T21** | **TFG** |
| --- | --- | --- |
| 2026-2045 | 1,650 (690 to 4,230) | 7,030 (3,510 to 12,600) |
| 2046-2065 | 3,020 (1,300 to 7,580) | 22,800 (13,000 to 37,300) |
| Total (40 years) | 4,680 (2,000 to 11,700) | 29,900 (16,500 to 50,100) |

BAU: business-as-usual; TFG: Tobacco free generation; T21: Tobacco-21.

**Supplementary Table 6. Deaths averted under each intervention compared to BAU, by timeframe.**

| **Timeframe** | **T21** | **TFG** |
| --- | --- | --- |
| 2026-2045 | 4 (2 to 9) | 17 (9 to 29) |
| 2046-2065 | 51 (24 to 122) | 442 (253 to 635) |
| Total (40 years) | 54 (25 to 130) | 459 (262 to 663) |

BAU: business-as-usual; TFG: Tobacco free generation; T21: Tobacco-21.

**Supplementary Table 7. Health impact under each intervention compared to BAU from 2025-2064: vape risk comparison.**

| **Outcome** | **T21** | | | | **TFG** | | | |
| --- | --- | --- | --- | --- | --- | --- | --- | --- |
|  | **Main analysis** | **Sensitivity analysis** | **Relative difference** | **Absolute difference** | **Main analysis** | **Sensitivity analysis** | **Relative difference** | **Absolute difference** |
| HALYs gained | 11,000 (4,710 to 27,600) | 11,000 (4,690 to 26,400) | 1 | 0 | 75,000 (41,900 to 124,000) | 60,700 (31,600 to 103,000) | 0.81 | -14,300 |
| Deaths averted | 54 (25 to 130) | 68 (-30 to 155) | 1.26 | 14 | 459 (262 to 663) | 383 (217 to 576) | 0.83 | -76 |

Main analysis: disease-risk from vaping set at mean 11% (95% uncertainty interval 5-20%) of the risk from smoking; sensitivity analysis: disease-risk from vaping set to 40% of that of smoking.

BAU: business-as-usual; TFG: Tobacco free generation; T21: Tobacco-21.

#### 3. Intervention parameterisation sensitivity analyses

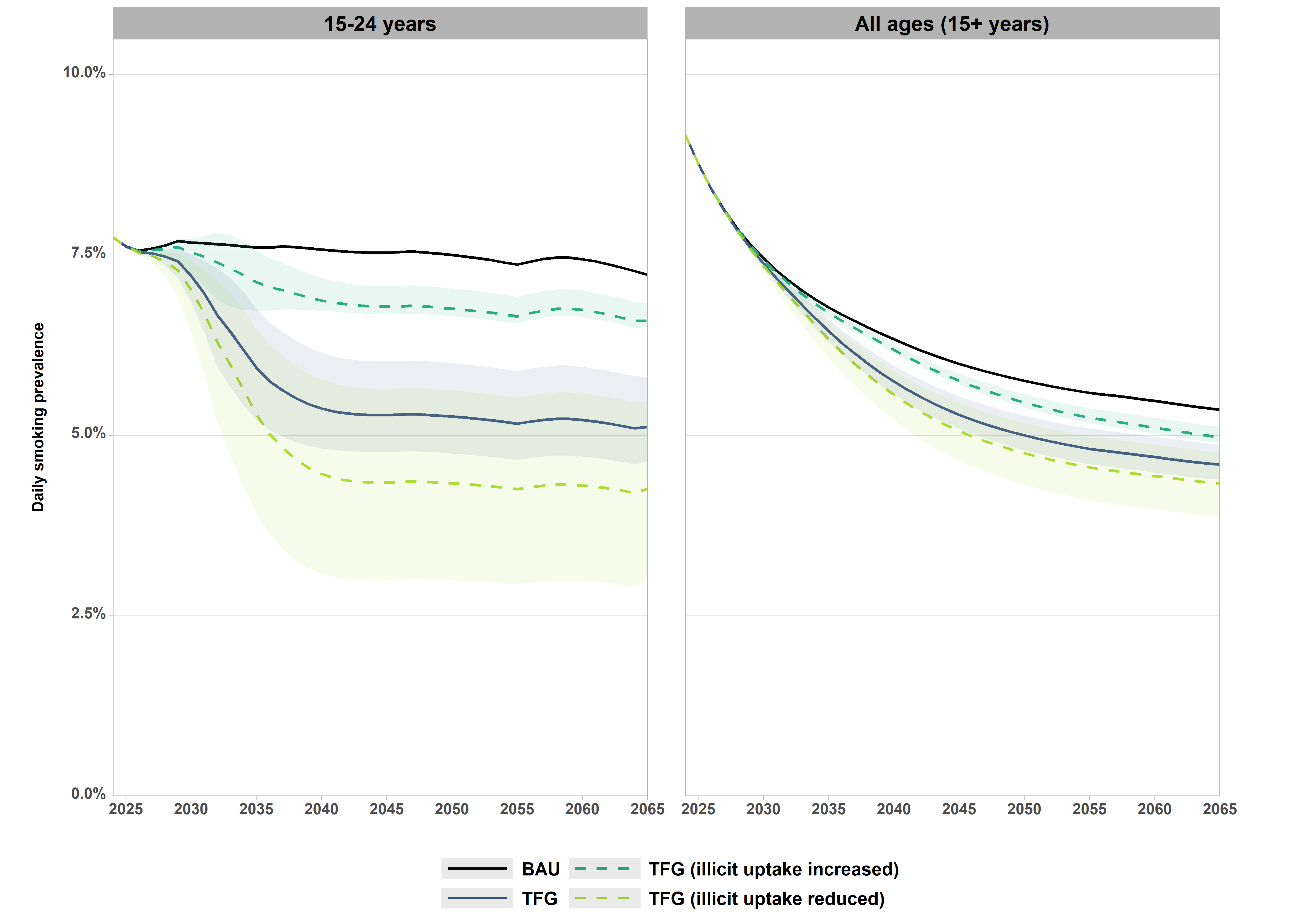

**Supplementary Figure 3. Daily smoking prevalence by age group and TFG sensitivity analysis scenario, 2025-2065.**

TFG initiated in 2026. Shaded area reflects the 95% uncertainty interval for the two policies (noting that there is no uncertainty around the BAU projections). Values shown in Supplementary Table 7. BAU: business-as-usual; TFG: Tobacco free generation.

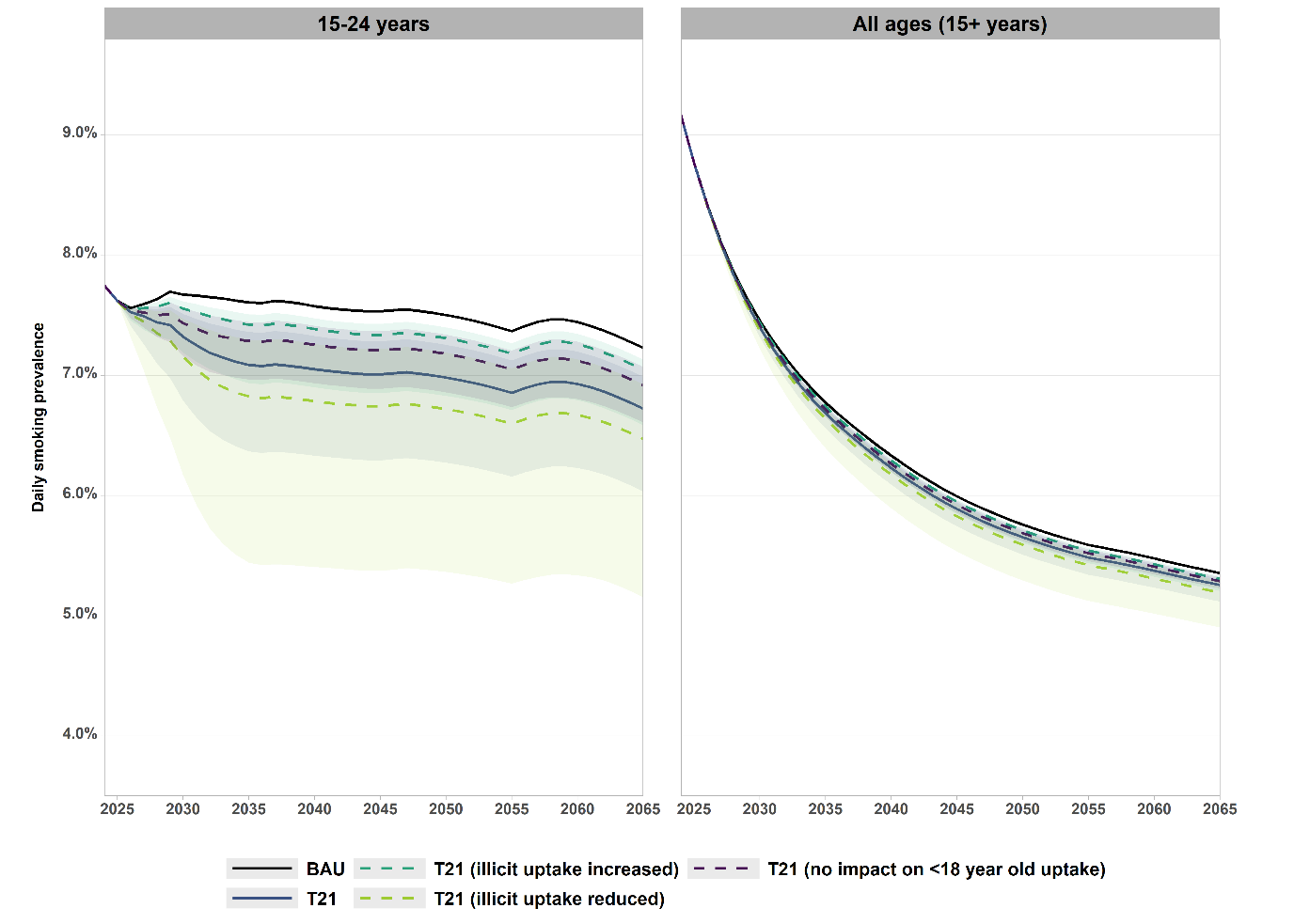

**Supplementary Figure 4. Daily smoking prevalence under T21 intervention, all ages and 15-24-year-olds, with varying intervention parameterisation.**

Note x-axis does not start at 0%. Values shown in Supplementary Table 7. BAU: business-as-usual; T21: Tobacco-21.

**Supplementary Table 8. Daily smoking prevalence under each intervention, with varying intervention parameterisation.**

| **Scenario** | **Age** | **2025*** | **2045** | **2065** |
| --- | --- | --- | --- | --- |
| **T21** | | | | |
| **T21 – main analysis** | **15-24** | 7.60% | 7% (6.3%-7.3%) | 6.7% (6%-7%) |
|  | **All (15+)** | 8.80% | 5.9% (5.7%-5.9%) | 5.3% (5.1%-5.3%) |
| **T21 – increase in illicit tobacco uptake** | **15-24** | 7.60% | 7.3% (6.9%-7.4%) | 7% (6.6%-7.1%) |
|  | **All (15+)** | 8.80% | 5.9% (5.8%-6%) | 5.3% (5.2%-5.3%) |
| **T21 – decrease in illicit tobacco uptake** | **15-24** | 7.60% | 6.7% (5.4%-7.2%) | 6.5% (5.2%-6.9%) |
|  | **All (15+)** | 8.80% | 5.8% (5.5%-5.9%) | 5.2% (4.9%-5.3%) |
| **T21 – no effect on <18-year-old uptake** | **15-24** | 7.60% | 7.2% (6.9%-7.4%) | 6.9% (6.6%-7.1%) |
|  | **All (15+)** | 8.80% | 5.9% (5.9%-6%) | 5.3% (5.2%-5.3%) |
| **TFG** | | | | |
| **TFG – main analysis** | **15-24** | 7.60% | 5.3% (4.8%-6%) | 5.1% (4.6%-5.8%) |
|  | **All (15+)** | 8.80% | 5.3% (5.1%-5.5%) | 4.6% (4.4%-4.9%) |
| **TFG – increase in illicit tobacco uptake** | **15-24** | 7.60% | 6.8% (6.7%-7.1%) | 6.6% (6.5%-6.8%) |
|  | **All (15+)** | 8.80% | 5.8% (5.7%-5.9%) | 5% (4.9%-5.1%) |
| **TFG – decrease in illicit tobacco uptake** | **15-24** | 7.60% | 4.3% (3%-5.6%) | 4.3% (3%-5.5%) |
|  | **All (15+)** | 8.80% | 5.1% (4.7%-5.4%) | 4.3% (3.9%-4.8%) |

*Pre-intervention: all values same, with no uncertainty.

BAU: business-as-usual; TFG: Tobacco free generation; T21: Tobacco-21.

**Supplementary Table 9. Health gain under each intervention compared to BAU, 2026-2065 (40-year time horizon), with varying intervention parameterisation.**

| **Scenario** | **Health-adjusted life years gained** | **Deaths averted** |
| --- | --- | --- |
| **T21** | | |
| **T21 – main analysis** | 11,000 (4,710 to 27,600) | 54 (25 to 130) |
| **T21 – increase in illicit tobacco uptake** | 4,260 (2,000 to 14,900) | 36 (17 to 97) |
| **T21 – decrease in illicit tobacco uptake** | 17,600 (6,520 to 51,400) | 97 (35 to 285) |
| **T21 – no effect on <18-year-old uptake** | 7,960 (3,560 to 17,500) | 44 (22 to 86) |
| **TFG** | | |
| **TFG – main analysis** | 75,000 (41,900 to 124,000) | 459 (262 to 663) |
| **TFG – increase in illicit tobacco uptake** | 26,900 (13,400 to 42,600) | 317 (181 to 455) |
| **TFG – decrease in illicit tobacco uptake** | 100,000 (50,500 to 168,000) | 611 (322 to 965) |

Health-adjusted life years (HALYs) are undiscounted.

BAU: business-as-usual; TFG: Tobacco free generation; T21: Tobacco-21.

#### 4. BAU parameterisation sensitivity analysis

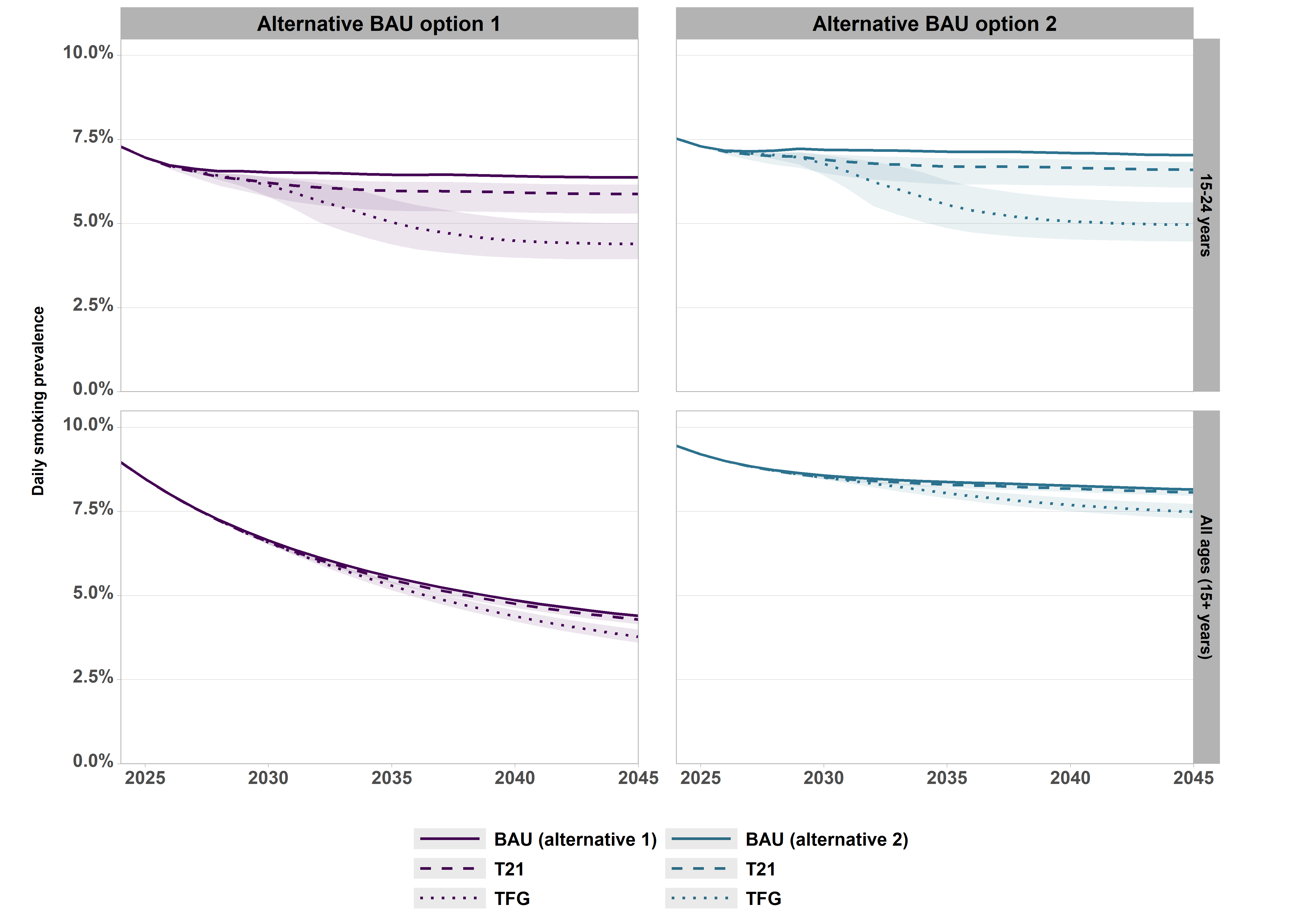

**Supplementary Figure 5. Daily smoking prevalence by age group and intervention, compared to two alternative BAU scenario parameterisations.** BAU alternative 1: lower rate of movement from vaping to smoking compared to main BAU; BAU alternative 2: lower rate of movement from smoking to vaping compared to main BAU. Details provided in Howe et al. [7]

**Supplementary Table 10. Relative and absolute change in daily smoking prevalence by 2045 under each intervention, comparing different BAU parameterisation.**

| **Intervention** | **Comparison** | **BAU parameterisation** | | |
| --- | --- | --- | --- | --- |
|  |  | **Main analysis BAU** | **Alternative BAU 1** | **Alternative BAU 2** |
| **T21** | **Absolute reduction** | 0.60pp | 0.44pp reduction | 0.49pp reduction |
|  | **Relative reduction** | 7.9% reduction | 6.2% reduction | 7.7% reduction |
| **TFG** | **Absolute reduction** | 2.3pp | 2.1pp reduction | 2.0pp reduction |
|  | **Relative reduction** | 30% reduction | 29% reduction | 31% reduction |

BAU: business-as-usual; TFG: Tobacco free generation; T21: Tobacco-21.

BAU alternative 1: lower rate of movement from vaping to smoking compared to main BAU; BAU alternative 2: lower rate of movement from smoking to vaping compared to main BAU. Details provided in Howe et al. [7]

#### 5. Policy impacts by illicit market enforcement level

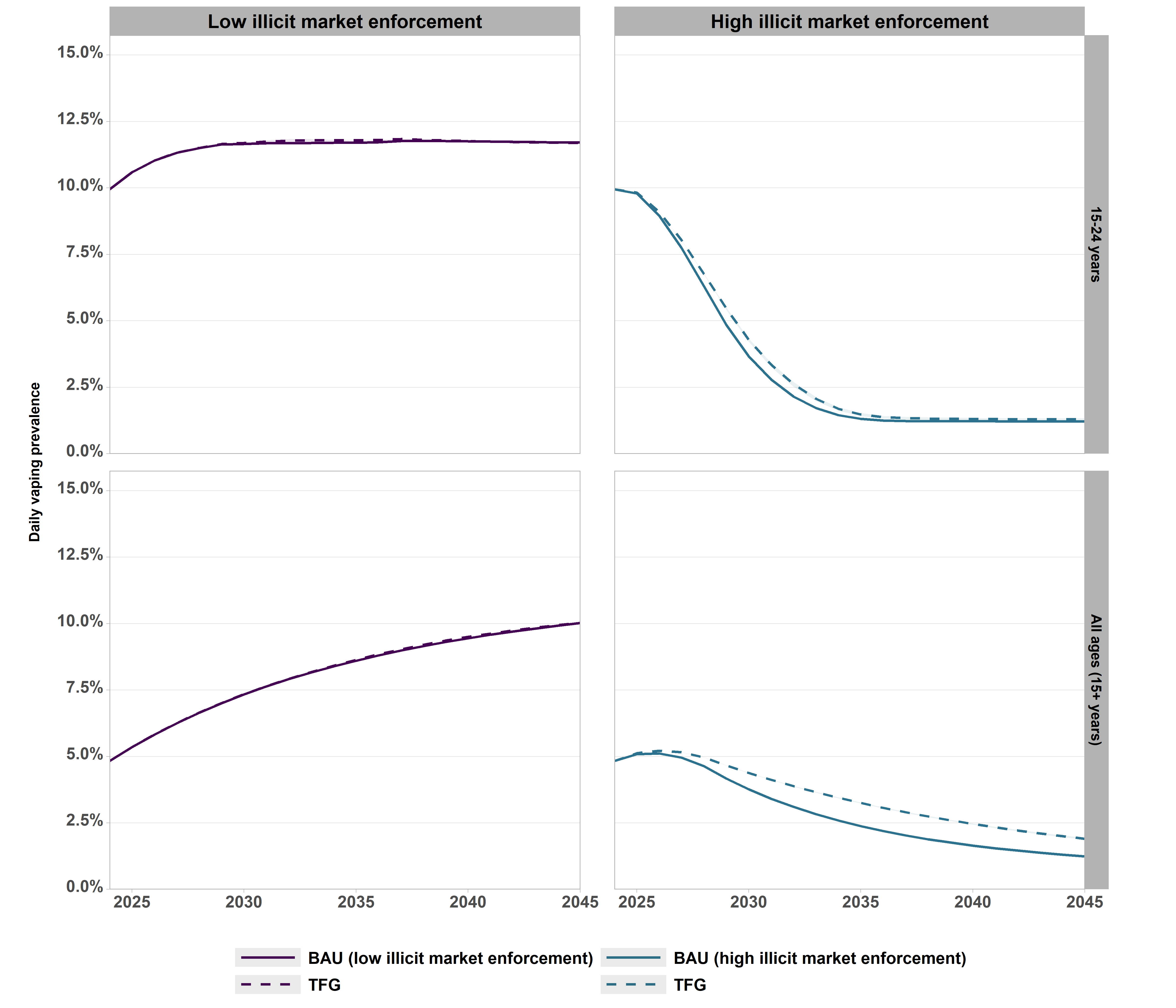

**Supplementary Figure 6. Daily vaping prevalence by age group, comparing TFG and background illicit market enforcement combinations.** TFG initiated in 2026. Shaded area reflects the 95% uncertainty interval for the two policies (noting that there is no uncertainty around the BAU projections or high illicit market enforcement scenario). The BAU line in the ‘high illicit market enforcement’ panels reflect increased illicit market enforcement only. BAU: business-as-usual; TFG: Tobacco free generation.

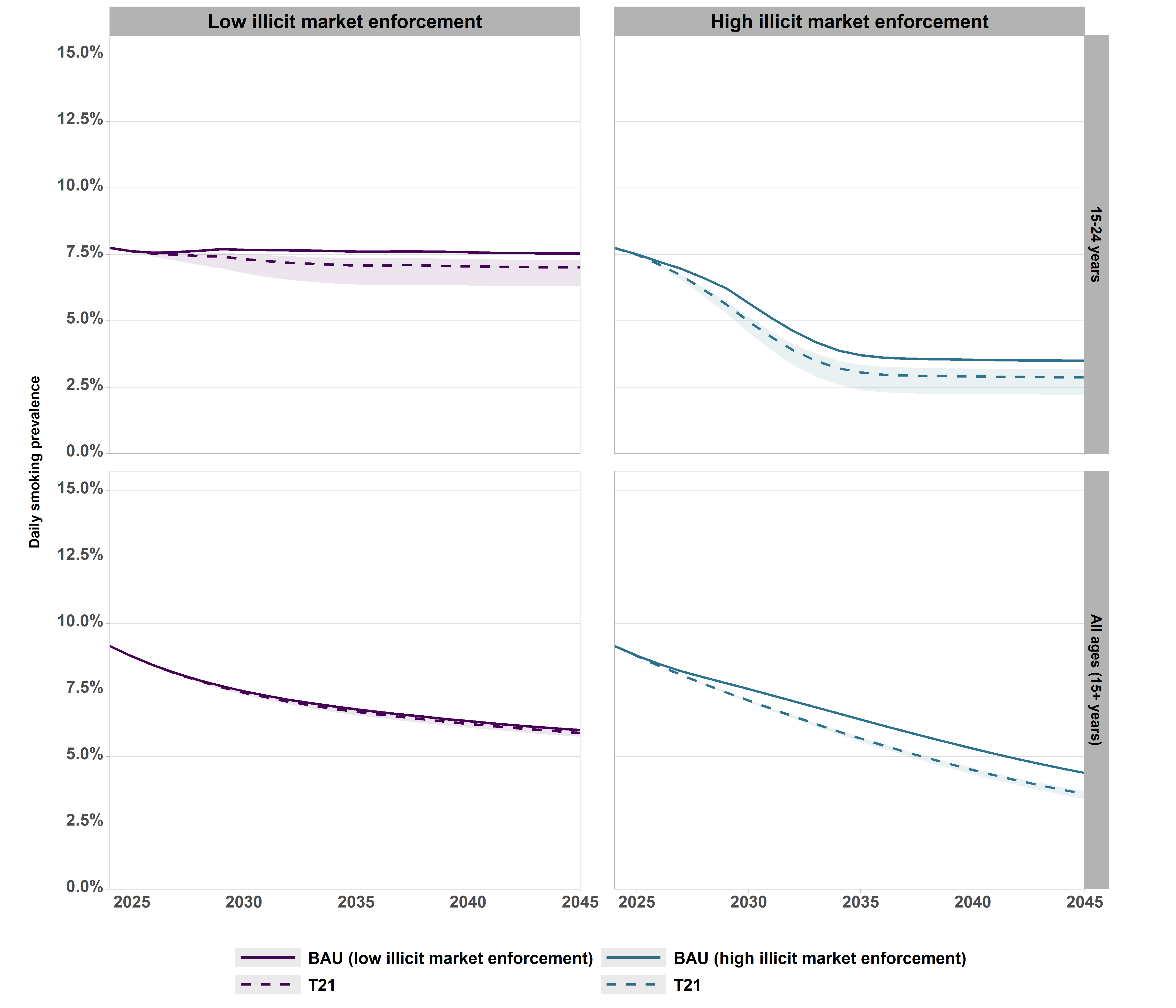

**Supplementary Figure 7. Daily smoking prevalence by age group, comparing T21 under varied background illicit market enforcement, 2025-2045.** TFG initiated in 2026. Shaded area reflects the 95% uncertainty interval for the policy (noting that there is no uncertainty around the BAU projections). Values shown in Supplementary Table 10. BAU: business-as-usual; TFG: Tobacco free generation.

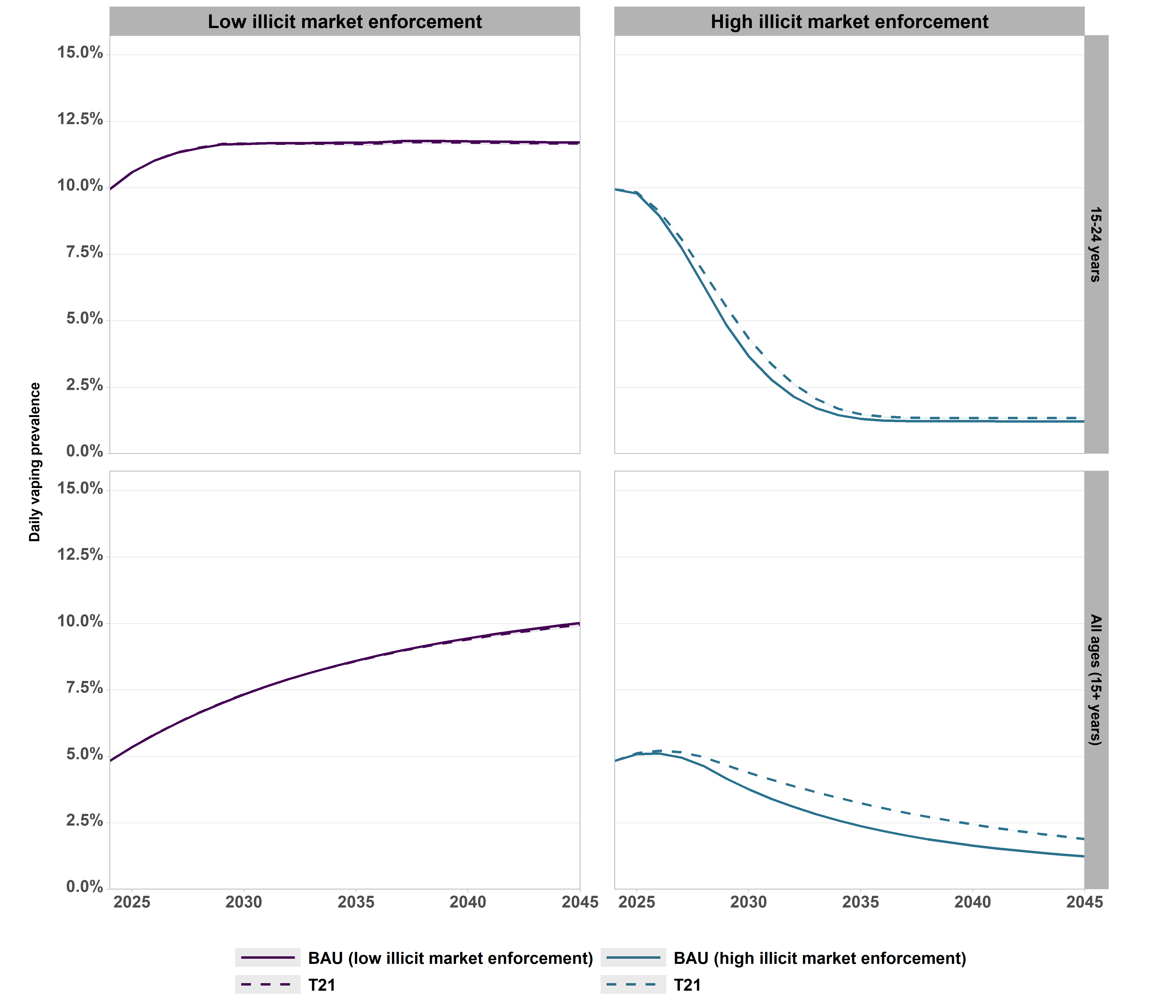

**Supplementary Figure 8. Daily vaping prevalence by age group, comparing T21 under varied background illicit market enforcement, 2025-2045.** TFG initiated in 2026. Shaded area reflects the 95% uncertainty interval for the policy (noting that there is no uncertainty around the BAU projections). Values shown in Supplementary Table 10. BAU: business-as-usual; TFG: Tobacco free generation.

**Supplementary Table 11. Daily smoking prevalence under each combined scenario, by age and year.**

| **Age group** | **2025*** | **2045** | **2065** |
| --- | --- | --- | --- |
| **BAU (High illicit market enforcement)^#^** | | | |
| **15+** | 8.80% | 4.4% | 2.58% |
| **15-24** | 7.50% | 3.5% | 3.33% |
| **25-34** | 11.50% | 5.2% | 4.93% |
| **35-44** | 10.80% | 8.1% | 3.67% |
| **45-54** | 10.80% | 5.7% | 2.35% |
| **55-64** | 9% | 3.6% | 2.41% |
| **65+** | 4.50% | 1.8% | 0.85% |
| **T21 + High enforcement** | | | |
| **15+** | 8.80% | 3.6% (3.4%-3.7%) | 2% (1.8%-2.2%) |
| **15-24** | 7.50% | 2.9% (2.2%-3.2%) | 2.7% (2.1%-3%) |
| **25-34** | 11.50% | 4.2% (3.6%-4.5%) | 4.1% (3.5%-4.3%) |
| **35-44** | 10.80% | 6.1% (6%-6.1%) | 2.9% (2.5%-3%) |
| **45-54** | 10.80% | 4.6% (4.6%-4.6%) | 1.8% (1.5%-1.9%) |
| **55-64** | 9% | 3.3% (3.3%-3.3%) | 1.7% (1.6%-1.7%) |
| **65+** | 4.50% | 1.8% (1.8%-1.8%) | 0.7% (0.7%-0.7%) |
| **TFG + High enforcement** | | | |
| **15+** | 8.80% | 3.1% (2.9%-3.3%) | 1.3% (1.1%-1.6%) |
| **15-24** | 7.50% | 1.3% (0.8%-2%) | 1.3% (0.9%-1.9%) |
| **25-34** | 11.50% | 2.6% (1.9%-3.4%) | 2% (1.5%-2.8%) |
| **35-44** | 10.80% | 5.9% (5.8%-6%) | 1.5% (1.2%-2%) |
| **45-54** | 10.80% | 4.6% (4.6%-4.6%) | 1.1% (0.9%-1.4%) |
| **55-64** | 9% | 3.3% (3.3%-3.3%) | 1.6% (1.6%-1.6%) |
| **65+** | 4.50% | 1.8% (1.8%-1.8%) | 0.7% (0.7%-0.7%) |

*Pre-intervention: all values same, with no uncertainty.

^#^No uncertainty under high illicit market enforcement alone.

BAU: business-as-usual; TFG: Tobacco free generation; T21: Tobacco-21.

**Supplementary Table 12. Relative and absolute difference in daily smoking prevalence in 2045 for each policy, under high vs. low enforcement.**

| **Scenario** | **T21** | | **TFG** | |
| --- | --- | --- | --- | --- |
|  | **Relative reduction** | **Absolute reduction** | **Relative reduction** | **Absolute reduction** |
| **15–24-year-olds** | | | | |
| **No increase in illicit market enforcement** | 7.9% | 0.6pp | 30.3% | 2.3pp |
| **High increase in illicit market enforcement** | 17.1% | 0.0pp | 62.9% | 2.2pp |
| **High vs. low illicit market enforcement** | 2.2x impact of T21 policy under high vs. low enforcement | Equal absolute impact of T21 policy under high vs. low enforcement | 2.1x impact of TFG policy under high vs. low enforcement | 0.1pp less impact of TFG policy under high vs. low enforcement |
| **All ages** | | | | |
| **No increase in illicit market enforcement** | 1.7% | 0.1pp | 11.7% | 0.7pp |
| **High increase in illicit market enforcement** | 18.2% | 0.8pp | 29.5% | 1.3pp |
| **High vs. low illicit market enforcement** | 3.8x impact of T21 policy under high vs. low enforcement | 0.7pp greater impact of T21 policy under high vs. low enforcement | 2.53x impact of TFG policy under high vs. low enforcement | 0.6pp greater impact of TFG policy under high vs. low enforcement |

TFG: Tobacco free generation; T21: Tobacco-21.

**Supplementary Table 13. Daily vaping prevalence under each combined scenario, by age and year.**

| **Age group** | **2025*** | **2045** | **2065** |
| --- | --- | --- | --- |
| **BAU (High illicit market enforcement)^#^** | | | |
| **15+** | 5.12% | 1.25% | 0.69% |
| **15-24** | 9.84% | 1.21% | 1.19% |
| **25-34** | 9.92% | 1.57% | 1.52% |
| **35-44** | 5.98% | 2.03% | 0.80% |
| **45-54** | 3.45% | 1.50% | 0.42% |
| **55-64** | 1.89% | 1.09% | 0.38% |
| **65+** | 0.60% | 0.55% | 0.30% |
| **T21 + High enforcement** | | | |
| **15+** | 5.12% | 1.90% (1.87%-1.91%) | 0.99% (0.95%-1.01%) |
| **15-24** | 9.84% | 1.34% (1.29%-1.36%) | 1.31% (1.26%-1.33%) |
| **25-34** | 9.92% | 2.08% (1.97%-2.14%) | 1.99% (1.88%-2.04%) |
| **35-44** | 5.98% | 3.83% (3.81%-3.85%) | 1.24% (1.16%-1.28%) |
| **45-54** | 3.45% | 2.61% (2.61%-2.61%) | 0.7% (0.65%-0.72%) |
| **55-64** | 1.89% | 1.62% (1.62%-1.62%) | 0.82% (0.81%-0.82%) |
| **65+** | 0.60% | 0.69% (0.69%-0.69%) | 0.51% (0.51%-0.51%) |
| **TFG + High enforcement** | | | |
| **15+** | 5.12% | 1.91% (1.87%-1.94%) | 0.96% (0.92%-1.01%) |
| **15-24** | 9.84% | 1.29% (1.26%-1.33%) | 1.27% (1.23%-1.3%) |
| **25-34** | 9.92% | 2.1% (1.96%-2.23%) | 1.93% (1.82%-2.06%) |
| **35-44** | 5.98% | 3.91% (3.86%-3.94%) | 1.16% (1.07%-1.25%) |
| **45-54** | 3.45% | 2.61% (2.61%-2.61%) | 0.66% (0.6%-0.72%) |
| **55-64** | 1.89% | 1.62% (1.62%-1.62%) | 0.83% (0.82%-0.84%) |
| **65+** | 0.60% | 0.69% (0.69%-0.69%) | 0.51% (0.51%-0.51%) |

*Pre-intervention: all values same, with no uncertainty.

^#^No uncertainty under high illicit market enforcement alone.

BAU: business-as-usual; TFG: Tobacco free generation; T21: Tobacco-21.

**Supplementary Table 14. Deaths averted by illicit market enforcement level, for T21 and TFG policies.**

| **Scenario** | **Timeframe** | **No policy** | **T21** | **TFG** |
| --- | --- | --- | --- | --- |
| **No increase in illicit market enforcement** | 2026-2045 | Reference | 4 (2 to 9) | 17 (9 to 29) |
|  | 2046-2065 | Reference | 51 (24 to 122) | 442 (253 to 635) |
|  | Total | Reference | 54 (25 to 130) | 459 (262 to 663) |
| **High increase in illicit market enforcement** | 2026-2045 | 7,220 (5,970 to 8,800) | 6,470 (5,450 to 7,710) | 6,480 (5,460 to 7,720) |
|  | 2046-2065 | 5,690 (4,120 to 7,690) | 14,400 (11,800 to 17,400) | 14,700 (12,200 to 17,800) |
|  | Total | 12,900 (10,400 to 16,200) | 20,800 (17,400 to 24,900) | 21,200 (17,700 to 25,400) |

*Negative deaths averted means more deaths occurred under the intervention scenario compared to BAU. This occurs because of a larger and older population under the intervention scenario (from fewer smoking-related deaths at younger ages in earlier years).

TFG: Tobacco free generation; T21: Tobacco-21.

**Supplementary Table 15. Health-adjusted life years gained under each intervention with varying illicit market enforcement, compared to BAU (Low enforcement), 2026-2065; discounted at 3% per annum.**

| **Scenario** | **T21** | **TFG** |
| --- | --- | --- |
| **Intervention alone** | 4,680 (2,000 to 11,700) | 29,900 (16,500 to 50,100) |
| **Intervention + High illicit market enforcement** | 339,000 (255,000 to 451,000) | 358,000 (267,000 to 476,000) |

TFG: Tobacco free generation; T21: Tobacco-21.

**Supplementary Table 16. Health impact under each intervention + high illicit market enforcement compared to BAU (Low enforcement), 2025-2064: vape risk comparison.**

| **Outcome** | **T21 + high illicit market enforcement** | | | | **TFG + high illicit market enforcement** | | | |
| --- | --- | --- | --- | --- | --- | --- | --- | --- |
|  | **Main analysis** | **Sensitivity analysis** | **Relative difference** | **Absolute difference** | **Main analysis** | **Sensitivity analysis** | **Relative difference** | **Absolute difference** |
| HALYs gained | 530,000 (378,000 to 738,000) | 768,000 (604,000 to 993,000) | 1.45 | 238,000 | 588,000 (416,000 to 824,000) | 822,000 (641,000 to 1,080,000) | 1.40 | 234,000 |
| Deaths averted | 8,200 (6,720 to 9,960) | 17,200 (14,700 to 20,100) | 2.10 | 9,000 | 8,630 (7,110 to 10,500) | 17,600 (15,100 to 20,700) | 2.04 | 8,970 |

Main analysis: disease-risk from vaping set at mean 11% (95% uncertainty interval 5-20%) of the risk from smoking; sensitivity analysis: disease-risk from vaping set to 40% of that of smoking.

BAU: business-as-usual; TFG: Tobacco free generation; T21: Tobacco-21.

#### 6. Policy impacts by SEIFA quintile

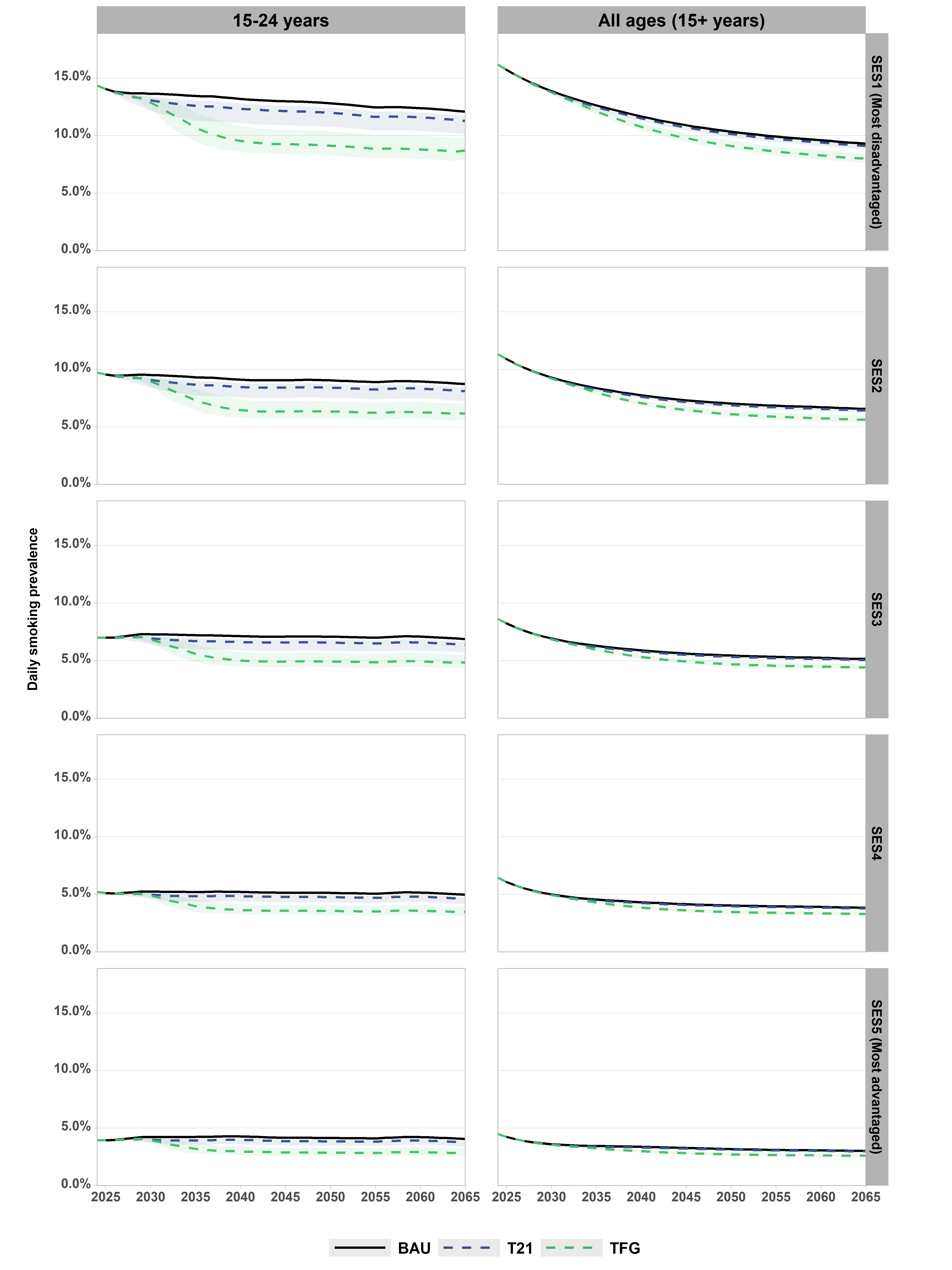

**Supplementary Figure 9. Daily smoking prevalence by SEIFA quintile and age, 2026-2065.** Values shown in Supplementary Table 14. BAU: business-as-usual; SES: socio-economic status; TFG: Tobacco free generation; T21: Tobacco-21.

**Supplementary Table 17. Daily smoking prevalence under each scenario, by SEIFA quintile.**

| **SEIFA** | **2025*** | **2045** | **2065** |
| --- | --- | --- | --- |
| **BAU (Low illicit market enforcement)^#^** | | | |
| **SES1** | 15.70% | 10.9% | 9.3% |
| **SES2** | 10.90% | 7.3% | 6.6% |
| **SES3** | 8.20% | 5.6% | 5.1% |
| **SES4** | 6.10% | 4.2% | 3.8% |
| **SES5** | 4.20% | 3.3% | 3% |
| **T21 only** | | | |
| **SES1** | 15.70% | 10.7% (10.4%-10.8%) | 9.1% (8.8%-9.2%) |
| **SES2** | 10.90% | 7.2% (7%-7.2%) | 6.4% (6.2%-6.5%) |
| **SES3** | 8.20% | 5.5% (5.4%-5.6%) | 5% (4.9%-5.1%) |
| **SES4** | 6.10% | 4.1% (4%-4.1%) | 3.8% (3.7%-3.8%) |
| **SES5** | 4.20% | 3.2% (3.2%-3.3%) | 3% (2.9%-3%) |
| **TFG only** | | | |
| **SES1** | 15.70% | 9.8% (9.5%-10.2%) | 8% (7.6%-8.5%) |
| **SES2** | 10.90% | 6.5% (6.2%-6.8%) | 5.6% (5.4%-6%) |
| **SES3** | 8.20% | 4.9% (4.7%-5.2%) | 4.4% (4.2%-4.7%) |
| **SES4** | 6.10% | 3.6% (3.5%-3.8%) | 3.3% (3.1%-3.5%) |
| **SES5** | 4.20% | 2.8% (2.7%-3%) | 2.6% (2.5%-2.8%) |
| **BAU (High illicit market enforcement)^#^** | | | |
| **SES1** | 15.70% | 8.9% | 5.5% |
| **SES2** | 10.90% | 5.6% | 3.4% |
| **SES3** | 8.20% | 4.0% | 2.3% |
| **SES4** | 6.10% | 2.7% | 1.5% |
| **SES5** | 4.30% | 1.9% | 1.0% |
| **T21 + High enforcement** | | | |
| **SES1** | 15.70% | 7.5% (7.2%-7.7%) | 4.4% (3.9%-4.7%) |
| **SES2** | 10.90% | 4.6% (4.3%-4.7%) | 2.6% (2.3%-2.8%) |
| **SES3** | 8.20% | 3.2% (3%-3.3%) | 1.8% (1.6%-1.9%) |
| **SES4** | 6.10% | 2.1% (2%-2.2%) | 1.1% (1%-1.2%) |
| **SES5** | 4.30% | 1.5% (1.4%-1.5%) | 0.8% (0.7%-0.8%) |
| **TFG + High enforcement** | | | |
| **SES1** | 15.70% | 6.6% (6.2%-7%) | 2.8% (2.3%-3.4%) |
| **SES2** | 10.90% | 4% (3.8%-4.3%) | 1.6% (1.4%-2%) |
| **SES3** | 8.20% | 2.8% (2.6%-3%) | 1.1% (0.9%-1.4%) |
| **SES4** | 6.10% | 1.8% (1.7%-1.9%) | 0.7% (0.6%-0.9%) |
| **SES5** | 4.30% | 1.3% (1.2%-1.4%) | 0.5% (0.4%-0.6%) |

*Pre-intervention: all values same, with no uncertainty.

^#^No uncertainty under BAU/high illicit market enforcement scenarios.

BAU: business-as-usual; SES: socio-economic status; TFG: Tobacco free generation; T21: Tobacco-21.

**Supplementary Table 18. Relative and absolute difference in daily smoking prevalence in 2045 for each policy vs. BAU (low enforcement), comparing SES1 vs. SES5 quintiles.**

| **Scenario** | **T21** | | **TFG** | |
| --- | --- | --- | --- | --- |
|  | **Relative reduction** | **Absolute reduction** | **Relative reduction** | **Absolute reduction** |
| **15–24-year-olds** | | | | |
| **SES1** | 6.9% | 0.8pp | 28.5% | 3.7pp |
| **SES5** | 7.1% | 0.3pp | 31.0% | 1.3pp |
| **SES1 vs. SES5** | 0.97x impact of T21 policy for SES1 vs. SES5 | 0.6pp greater impact of T21 policy for SES1 vs. SES5 | 0.92x impact of TFG policy for SES1 vs. SES5 | 2.4pp greater impact of TFG policy for SES1 vs. SES5 |
| **All ages** | | | | |
| **SES1** | 1.8% | 0.2pp | 10.1% | 1.1pp |
| **SES5** | 3.0% | 0.1pp | 15.2% | 0.5pp |
| **SES1 vs. SES5** | 0.61x impact of T21 policy for SES1 vs. SES5 | 0.1pp greater impact of T21 policy for SES1 vs. SES5 | 0.67x impact of TFG policy for SES1 vs. SES5 | 0.6pp greater impact of TFG policy for SES1 vs. SES5 |

SES: socio-economic status; TFG: Tobacco free generation; T21: Tobacco-21.

**Supplementary Table 19. Relative and absolute difference in daily smoking prevalence in 2045 for each policy vs. BAU (high enforcement), comparing SES1 vs. SES5 quintiles.**

| **Scenario** | **T21 + high illicit market enforcement** | | **TFG + high illicit market enforcement** | |
| --- | --- | --- | --- | --- |
|  | **Relative reduction** | **Absolute reduction** | **Relative reduction** | **Absolute reduction** |
| **15–24-year-olds** | | | | |
| **SES1** | 16.4% | 0.34pp | 61.3% | 3.7pp |
| **SES5** | 19.5% | 1.04pp | 62.0% | 1.3pp |
| **SES1 vs. SES5** | 0.84x impact of T21 policy for SES1 vs. SES5 | 0.71pp greater impact of T21 policy for SES1 vs. SES5 | 0.99x impact of TFG policy for SES1 vs. SES5 | 2.4pp greater impact of TFG policy for SES1 vs. SES5 |
| **All ages** | | | | |
| **SES1** | 15.7% | 1.4pp | 26.5% | 2.4pp |
| **SES5** | 20.0% | 0.4pp | 32.4% | 1.6pp |
| **SES1 vs. SES5** | 0.78x impact of T21 policy for SES1 vs. SES5 | 1.0pp greater impact of T21 policy for SES1 vs. SES5 | 0.82x impact of TFG policy for SES1 vs. SES5 | 1.8pp greater impact of TFG policy for SES1 vs. SES5 |

SES: socio-economic status; TFG: Tobacco free generation; T21: Tobacco-21.

**Supplementary Table 20. Health gain under each intervention compared to BAU, by SEIFA quintile, 2026-2065.**

| **Scenario** | **SEIFA quintile** | **Absolute HALY gain** | **Age-standardised HALY gain per 100,000 person-years*** | **Relative difference in age-standardised health gain by quintile** |
| --- | --- | --- | --- | --- |
| **T21** | **SES1 (most disadvantaged)** | 3,300 (1,420 to 8,520) | 1.5 (0.7 to 4.0) | 3.8 |
|  | **SES2** | 2,540 (1,090 to 6,480) | 1.1 (0.5 to 2.8) | 2.8 |
|  | **SES3** | 2,360 (1,010 to 5,920) | 0.9 (0.4 to 2.2) | 2.2 |
|  | **SES4** | 1,580 (687 to 3,850) | 0.6 (0.3 to 1.4) | 1.5 |
|  | **SES5 (most advantaged)** | 1,150 (505 to 2,760) | 0.4 (0.2 to 1.0) | Reference |
| **TFG** | **SES1 (most disadvantaged)** | 19,200 (10,700 to 31,500) | 9.0 (5.0 to 15) | 2.3 |
|  | **SES2** | 15,700 (8,730 to 26,000) | 6.9 (3.8 to 11) | 1.7 |
|  | **SES3** | 15,900 (8,910 to 26,500) | 5.9 (3.3 to 9.8) | 1.5 |
|  | **SES4** | 13,400 (7,520 to 22,300) | 4.9 (2.8 to 8.2) | 1.2 |
|  | **SES5 (most advantaged)** | 10,800 (6,070 to 17,900) | 4.0 (2.2 to 6.6) | Reference |

*Age-standardised to the 2001 Australian population, per (28)

HALYs are undiscounted.

HALY: health-adjusted life year; SES: socio-economic status; TFG: Tobacco free generation; T21: Tobacco-21.
